# Neutralizing antibody responses to SARS-CoV-2 variants in vaccinated Ontario long-term care home residents and workers

**DOI:** 10.1101/2021.08.06.21261721

**Authors:** Kento T. Abe, Queenie Hu, Mohammad Mozafarihashjin, Reuben Samson, Kathy Manguiat, Alyssia Robinson, Bhavisha Rathod, W. Rod Hardy, Jenny H. Wang, Mariam Iskilova, Adrian Pasculescu, Mahya Fazel-Zarandi, Angel Li, Aimee Paterson, Gary Chao, Karen Green, Lois Gilbert, Shiva Barati, Nazrana Haq, Alyson Takaoka, Julia Garnham Takaoka, Keelia Quinn De Launay, Christine Fahim, Salma Sheikh-Mohamed, Yuko Arita, Yves Durocher, Eric G. Marcusson, Jennifer L. Gommerman, Mario Ostrowski, Karen Colwill, Sharon E. Straus, Heidi Wood, Allison J. McGeer, Anne-Claude Gingras

**Affiliations:** Lunenfeld-Tanenbaum Research Institute at Mount Sinai Hospital, Sinai Health, Toronto, ON, Canada; Department of Molecular Genetics, University of Toronto, Toronto, ON, Canada; Department of Microbiology, at Mount Sinai Hospital, Sinai Health System, Toronto, ON, Canada; National Microbiology Laboratory, Public Health Agency of Canada, Winnipeg, MN, Canada; Department of Immunology, University of Toronto, Toronto, ON, Canada; Li Ka Shing Knowledge Institute, St. Michael’s Hospital, Unity Health Toronto, Toronto, ON, Canada; Providence Therapeutics Holdings, Inc.; Calgary, Alberta, Canada; Mammalian Cell Expression, Human Health Therapeutics Research Centre, National Research Council Canada, Montréal, QC, Canada; Marcusson Consulting, San Francisco, CA, USA; Department of Medicine, University of Toronto, Toronto, ON, Canada; Institute of Health Policy, Management and Evaluation, University of Toronto, Toronto, ON, Canada

## Abstract

Prioritizing Ontario’s long-term care home (LTCH) residents for vaccination against severe acute respiratory syndrome coronavirus 2 has drastically reduced their disease burden; however, recent LTCH outbreaks of variants of concern (VOCs) have raised questions regarding their immune responses. In 198 residents, mRNA vaccine dose 1 elicited partial spike and receptor binding domain antibody responses, while the second elicited a response at least equivalent to convalescent individuals in most residents. Residents administered mRNA-1273 (Moderna) mounted stronger total and neutralizing antibody responses than those administered BNT162b2 (Pfizer-BioNTech). Two to four weeks after dose 2, residents (*n* = 119, median age 88) produced 4.8–6.3-fold fewer neutralizing antibodies than staff (*n* = 78; median age 47) against wild-type (with D614G) pseudotyped lentivirus, and residents administered BNT162b2 produced 3.89-fold fewer neutralizing antibodies than those who received mRNA-1273. These effects were exacerbated upon serum challenge with pseudotyped VOC spike, with up to 7.94-fold reductions in B.1.351 (Beta) neutralization. Cumulatively, weaker vaccine stimulation, age/comorbidities, and the VOC produced an ∼130-fold reduction in apparent neutralization titers in LTCH residents and 37.9% of BNT162b2-vaccinated residents had undetectable neutralizing antibodies to B.1.351. Continued immune response surveillance and additional vaccine doses may be required in this population with known vulnerabilities.

## Introduction

The first waves of the coronavirus disease 2019 (COVID-19) pandemic disproportionately affected older adults, particularly in long-term care homes (LTCH) worldwide. In Canada, LTCH residents accounted for up to 85% of COVID-19 deaths in the first year of the pandemic [1]. Prioritized vaccination has drastically decreased LTCH outbreaks and associated mortality [2]. In Ontario, vaccination began mid-December 2020, with hospital-based healthcare workers and LTCH staff among the first to receive the BNT162b2 (Pfizer-BioNTech) vaccine. LTCH residents in Ontario primarily received the mRNA-1273 (Moderna) vaccine, which was initially thought to be easier to transport to LTCH settings [2]; however, some LTCHs used BNT162b2 to vaccinate both staff and residents. Despite the overwhelming success of vaccination (combined with other public health measures) in decreasing mortality and infection in these settings, LTCH outbreaks have recently re-emerged, resulting in severe disease and death in some residents. Outbreaks with more severe disease have been associated with immune-escape variants, such as P.1 (Gamma)[3-7].

Several recent reports (primarily using the BNT162b2 vaccine) have described weaker humoral responses (in several cases including neutralizing antibody titers) in LTCH residents than in control groups, after both one and two vaccine doses [8, 9], with a particularly poor humoral response after the first dose in individuals not previously infected with severe acute respiratory syndrome coronavirus 2 (SARS-CoV-2). Other studies have demonstrated strong age-related declines in seroconversion after the first dose of BNT162b2 [9-11]. This weaker and less durable response likely contributes to the reduced vaccine effectiveness in LTCH residents and health care workers reported in some studies [12], and to recent LTCH outbreaks as well. Notably, in both transplant recipients [13, 14] and healthy individuals [15], the mRNA-1273 vaccine induced a stronger humoral response than BNT162b2; however, whether these differences exist between the mRNA vaccines used in LTCH settings (BNT162b2 vs mRNA-1273) remains unknown. The kinetics of antibody production and decline, and crucially, the ability of these antibodies to neutralize variants of concern (VOCs) effectively, also remain unreported in this population with known vulnerabilities.

While the best correlates for immune protection are yet to be determined, a recent study attempting to reconcile all clinical trials by normalizing neutralization titers across them revealed a clear correlation between this value and vaccine effectiveness [16]. This suggests that neutralization is a good proxy for overall vaccine effectiveness, as previously demonstrated for influenza [17]. While complete vaccination in healthy individuals still induces cross-neutralization of all currently circulating VOCs, effective VOC neutralization titers are often reduced several-fold [18-20]. This is also consistent with reports that breakthrough infections and LTCH outbreaks are enriched for immune-escape variants such as B.1.351 (Beta) [21] [4]. It is noteworthy that in one outbreak investigation, neutralization titers in individuals that became infected were lower than those in non-infected individuals [4]. The factors mediating immune escape, including biological factors (i.e., immune response variations between individuals) and mutations in the variants themselves, remain unknown; however, recent studies have begun to address these questions (e.g., [22]).

Previously, we developed ELISAs to detect IgG, IgA, and IgM levels against the SARS-CoV-2 spike trimer, spike receptor binding domain (RBD), and nucleocapsid protein (NP), which we used to monitor the humoral responses of individuals in the first 4 months post-infection [23]. A spike-pseudotyped lentiviral assay [24] was also optimized and used to monitor neutralization in cohorts of plasma donors in parallel to plaque reduction neutralization titer (PRNT) assays and a surrogate neutralization ELISA [25]. This assay has since been modified to include spike sequences from the B.1.1.7 (Alpha), B.1.351 (Beta), P.1 (Gamma) and B.1.617.2 (Delta) VOCs [26].

Here, we evaluate humoral responses to vaccination in cohorts of LTCH residents after one or two doses of mRNA-1273 or BNT162b2. We also compare the responses in LTCH residents and staff at 2–4 weeks post-dose 2, examining both anti-spike and anti-RBD IgGs, and neutralizing antibodies to wild-type spike as well as the B.1.1.7, B.1.351, P.1 and B.1.617.2 variants. The results reveal deficiencies in the humoral immune responses of LTCH residents that may leave them vulnerable to infection due to VOCs.

## Results

### A robust antibody response in LTCH residents requires two doses of mRNA vaccine

We enrolled 198 residents of 4 long term care homes in south-central Ontario; residents in three of these homes received mRNA-1273 and those in the fourth received BNT162b2 (Table 1). All vaccines were delivered on manufacturer’s recommended schedules in January and February of 2021.

**Table 1:**
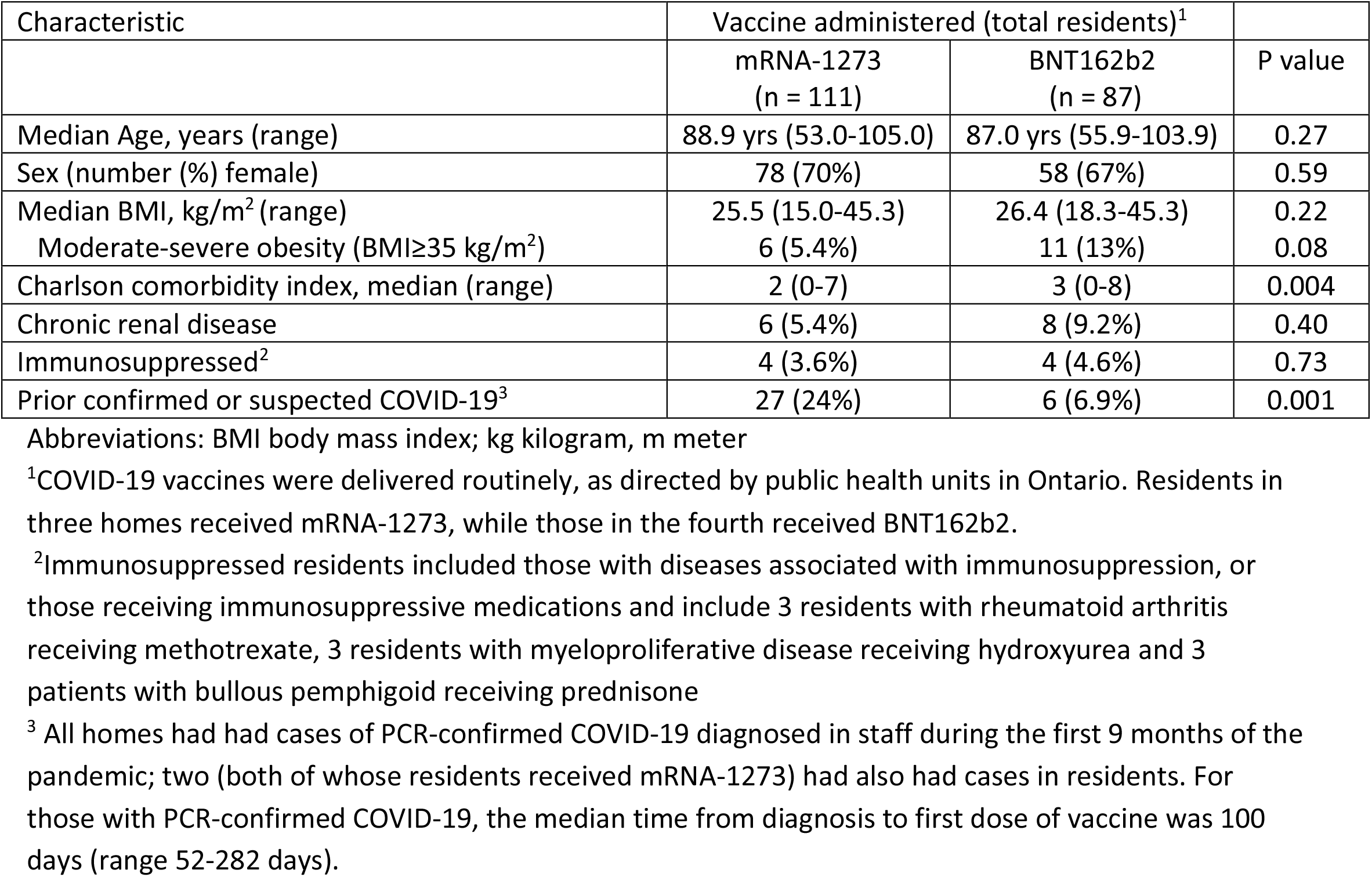
Characteristics of residents participating in COVID-19 vaccine immunogenicity study, Ontario, 2021

| Characteristic | Vaccine administered (total residents)<sup>1</sup> |  | P value |
| --- | --- | --- | --- |
|  | mRNA-1273<br>(n = 111) | BNT162b2<br>(n = 87) |  |
| Median Age, years (range) | 88.9 yrs (53.0-105.0) | 87.0 yrs (55.9-103.9) | 0.27 |
| Sex (number (%) female) | 78 (70%) | 58 (67%) | 0.59 |
| Median BMI, kg/m<sup>2</sup> (range) | 25.5 (15.0-45.3) | 26.4 (18.3-45.3) | 0.22 |
| Moderate-severe obesity (BMI≥35 kg/m<sup>2</sup>) | 6 (5.4%) | 11 (13%) | 0.08 |
| Charlson comorbidity index, median (range) | 2 (0-7) | 3 (0-8) | 0.004 |
| Chronic renal disease | 6 (5.4%) | 8 (9.2%) | 0.40 |
| Immunosuppressed<sup>2</sup> | 4 (3.6%) | 4 (4.6%) | 0.73 |
| Prior confirmed or suspected COVID-19<sup>3</sup> | 27 (24%) | 6 (6.9%) | 0.001 |

Abbreviations: BMI body mass index; kg kilogram, m meter
<sup>1</sup>COVID-19 vaccines were delivered routinely, as directed by public health units in Ontario. Residents in three homes received mRNA-1273, while those in the fourth received BNT162b2.
<sup>2</sup>Immunosuppressed residents included those with diseases associated with immunosuppression, or those receiving immunosuppressive medications and include 3 residents with rheumatoid arthritis receiving methotrexate, 3 residents with myeloproliferative disease receiving hydroxyurea and 3 patients with bullous pemphigoid receiving prednisone
<sup>3</sup> All homes had had cases of PCR-confirmed COVID-19 diagnosed in staff during the first 9 months of the pandemic; two (both of whose residents received mRNA-1273) had also had cases in residents. For those with PCR-confirmed COVID-19, the median time from diagnosis to first dose of vaccine was 100 days (range 52-282 days).

We obtained samples (dried blood spot (DBS) or serum; see the Methods and figure legends for details) from participating residents prior to or up to 4 days after first dose of vaccine (baseline), immediately prior to their second dose of the same vaccine (post-dose 1) and 2–4 weeks after a second dose of the same vaccine (Figure 1A). Samples were analyzed by automated ELISA and all values are reported in relation to a synthetic standard that enables assay calibration (see the Methods for details).

**Figure 1.**
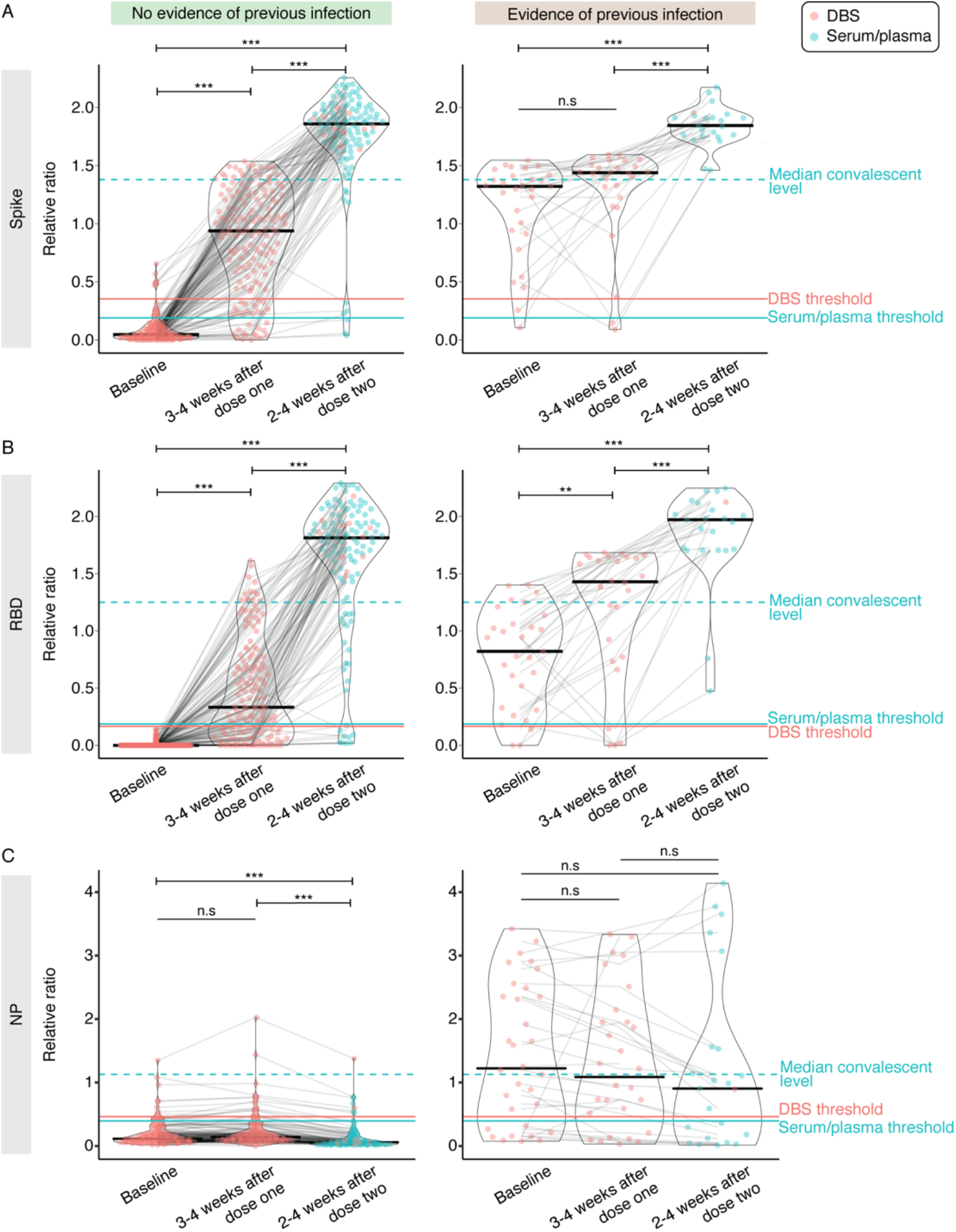
IgG responses to the first and second doses of mRNA vaccine in LTCH residents. A–C) Raw chemiluminescent ELISA reads were normalized to a synthetic standard curve to determine relative ratios for spike, spike RBD, and NP at three timepoints. DBS or serum/plasma samples were taken at before or up to 2 weeks after the first dose (baseline; *n* = 174 for no previous infection and *n* = 33 for previous infection), 3–4 weeks after the first dose (*n* = 157 for no previous infection and *n* = 29 for previous infection) or 2–4 weeks after the second dose (*n* = 130 for no previous infection and *n* = 23 for previous infection). Violin plots show the relative ratio distributions at each timepoint. Turquoise and salmon points are results from serum/plasma (0.0625 μL sample used) and ratio-corrected DBS eluate (2.5 µl sample used), respectively (see Methods). Solid black lines indicate the median ratio value for each violin, and grey lines connect data points for each participant. The solid turquoise and salmon lines are the seropositivity thresholds for the serum/plasma and DBS samples, respectively. The dashed turquoise line is the median value of 211 convalescent COVID-19 patients. Plots are faceted based on whether patients had evidence of previous infection as determined by serology (samples positive for two or more antigen tests at baseline) and/or qPCR. The Kruskal-Wallis test and Dunn’s post test was used to determine significance. P-values were adjusted using Bonferroni correction. **** P < 0.001, * P < 0.05.

Overall, 25 residents had previous (q)PCR-confirmed SARS-CoV-2 infections and an additional 8 residents who scored positive in two or more of the ELISA assays (against the spike trimer, spike RBD, and NP), were considered to have evidence of prior infection (Table 2).

**Table 2:**
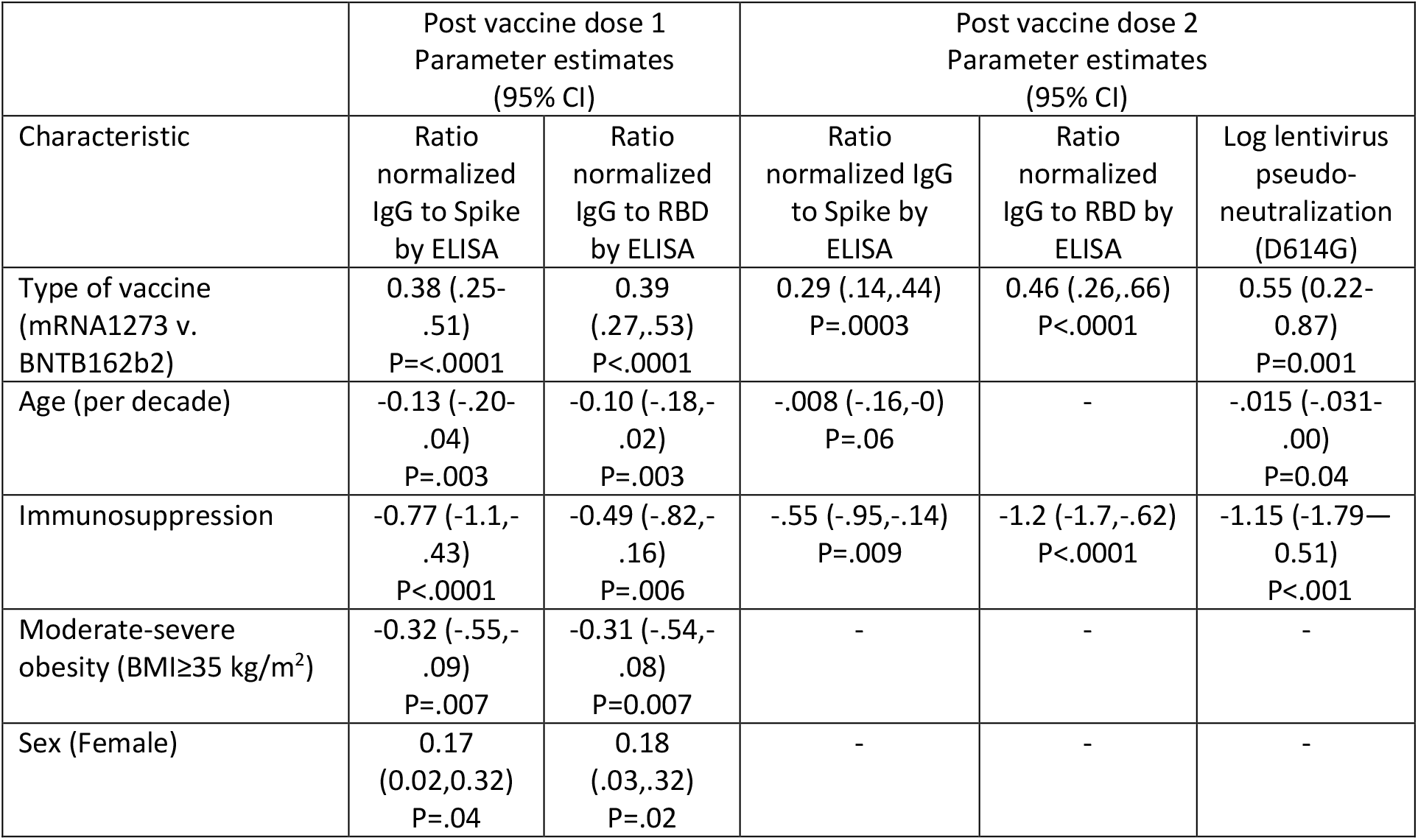
Parameter estimates in multivariable analysis of factors associated with spike, RBD and lentivirus pseudoneutralization response

| Characteristic | Post vaccine dose 1<br>Parameter estimates<br>(95% CI) |  | Post vaccine dose 2<br>Parameter estimates<br>(95% CI) |  |  |
| --- | --- | --- | --- | --- | --- |
|  | Ratio<br>normalized<br>IgG to Spike<br>by ELISA | Ratio<br>normalized<br>IgG to RBD<br>by ELISA | Ratio<br>normalized IgG<br>to Spike by<br>ELISA | Ratio<br>normalized<br>IgG to RBD by<br>ELISA | Log lentivirus<br>pseudo-<br>neutralization<br>(D614G) |
| Type of vaccine<br>(mRNA1273 v.<br>BNTB162b2) | 0.38 (.25-<br>.51)<br>P<.0001 | 0.39<br>(.27,.53)<br>P<.0001 | 0.29 (.14,.44)<br>P=.0003 | 0.46 (.26,.66)<br>P<.0001 | 0.55 (0.22-0.87)<br>P=0.001 |
| Age (per decade) | -0.13 (-.20-<br>.04)<br>P=.003 | -0.10 (-.18,-<br>.02)<br>P=.003 | -.008 (-.16,-0)<br>P=.06 | - | -.015 (-.031-<br>.00)<br>P=0.04 |
| Immunosuppression | -0.77 (-1.1,-<br>.43)<br>P<.0001 | -0.49 (-.82,-<br>.16)<br>P=.006 | -.55 (-.95,-.14)<br>P=.009 | -1.2 (-1.7,-.62)<br>P<.0001 | -1.15 (-1.79—<br>0.51)<br>P<.001 |
| Moderate-severe<br>obesity (BMI≥35 kg/m<sup>2</sup>) | -0.32 (-.55,-<br>.09)<br>P=.007 | -0.31 (-.54,-<br>.08)<br>P=0.007 | - | - | - |
| Sex (Female) | 0.17<br>(0.02,0.32)<br>P=.04 | 0.18<br>(.03,.32)<br>P=.02 | - | - | - |

Among those without evidence of previous infection, the first dose elicited seroconversion for both spike and RBD IgGs in 66.4% (95/143) of LTCH residents, and seroconversion for either IgG in 79.7% (114/143; Figure 1A, B; Supplemental Table 1). In 11.9% (17/143) of residents, anti-spike IgG levels were greater than the median levels seen in a group of convalescent patients who recovered from COVID-19, and 7.7% (11/143) of residents had higher levels of anti-RBD antibodies. At 2–4 weeks post-dose two, 92.2% (106/130) of residents had seroconverted for both spike and its RBD (97.4% spike; 92.2% RBD). The relative ratio was greater than the medians of the convalescents, at 91.3% for spike IgG (105/115) and 80.0% for RBD IgG (92/115), respectively. At both points, there was a wide range of response to vaccination. The same overall trends were detected in previously infected residents, with both doses eliciting higher levels of spike and RBD antibodies (Figure 1A, B).

As expected, since the vaccines do not contain NP sequences, levels of anti-NP antibodies did not increase across the timepoints regardless of previous infection, and there was a relative decrease signal at later time points in the cohort with evidence of previous infection that may be related to waning antibodies (Figure 1C). In summary, the vaccines elicit stronger anti-spike and anti-RBD IgG responses in older individuals 2–4 weeks post-dose 2 than after the first dose.

### The mRNA-1273 vaccine elicits a stronger humoral response than the BNT162b2 vaccine in both LTCH residents and staff

The variability in responses in residents that were not previously infected—particularly to dose 1— prompted us to explore the reasons underlying it.

In bivariable analyses of factors associated with spike and RBD responses at after dose 1 of vaccine in residents who had not previously had PCR-confirmed COVID-19, younger age, lower Charlson score, absence of immunosuppression and chronic renal disease, BMI <35 kg/m^2^, and type of vaccine (mRNA-1273 vs. BNT162b2) were associated with stronger responses to the first dose of vaccine. Trends were similar for response after the second dose of vaccine, although the associations with BMI, age and Charlson index were no longer significant. In multivariable analysis, type of vaccine, age, BMI, immunosuppression and sex were associates with spike and RBD response to dose one, but only type of vaccine and immunosuppression remained significantly associated with the response to dose 2 (Table 2).

This difference in response by vaccine type was consistent across responses to both spike and its RBD, and was particularly notable after dose 1, though it remained statistically significant at 2–4 post-dose 2 (Figure 2A, B). The differences were noted both when the assay was performed in a mode optimized for seropositivity detection and after 4-fold dilution of the samples to better capture differential vaccine responses (Supplemental Figure 1A, B).

**Figure 2.**
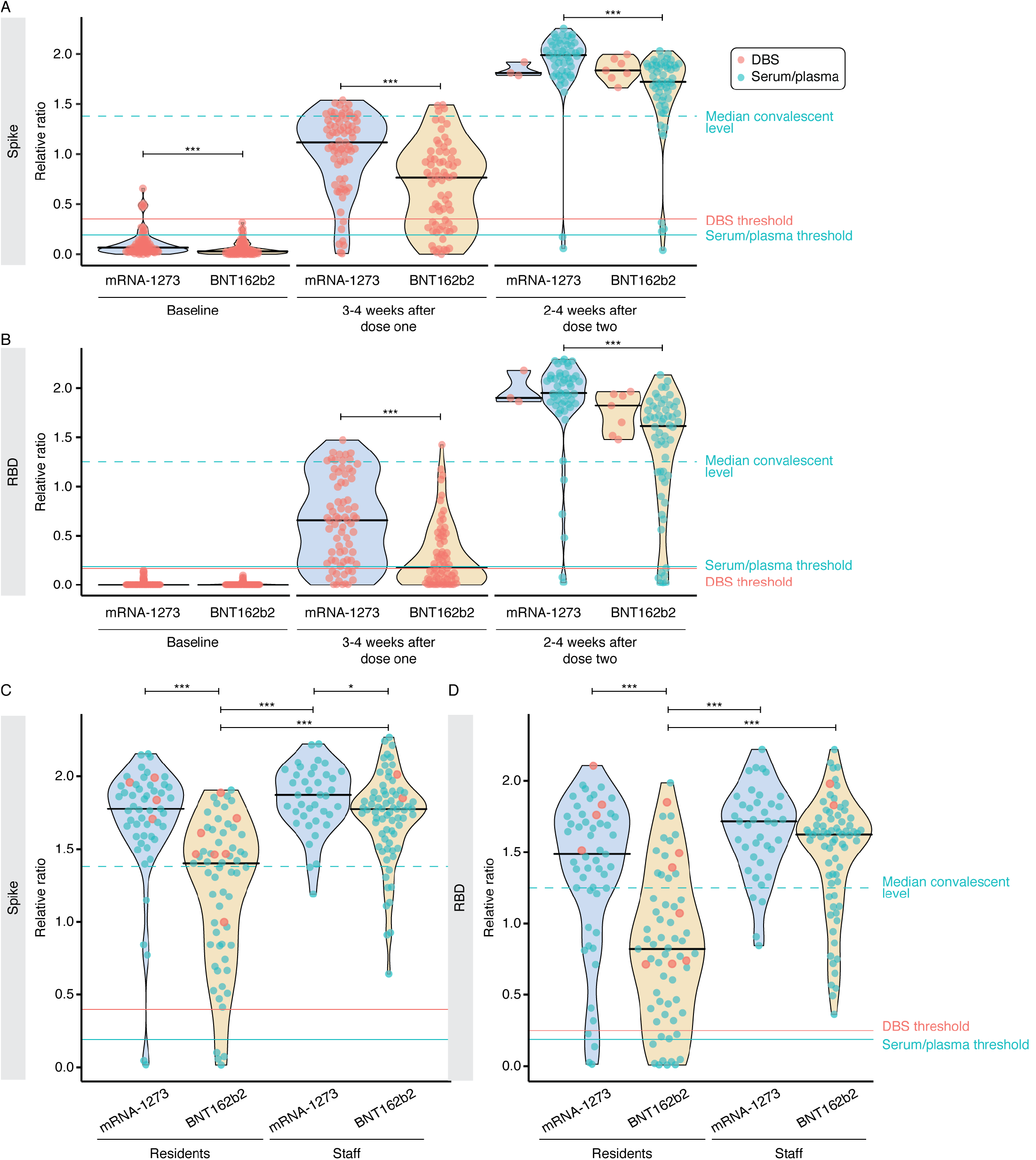
Contributions of vaccine type to response variability in vaccinated individuals. A–B) Levels of serum anti-spike and anti-RBD IgG levels stratified by vaccine type (mRNA-1273, Moderna; BNT162b2, Pfizer) at each time point in not previously infected individuals. Solid black lines indicate the median ratio values for each violin. Turquoise and salmon points are results from serum/plasma (0.0625 μL sample used) and ratio-corrected DBS eluate (2.5 µl sample used), respectively. The dashed turquoise line is the median value of 211 convalescent COVID-19 patients. The Mann-Whitney U test was used to compare between vaccine types at each timepoint, and P-values were adjusted using Bonferroni correction. At baseline, 3-4 weeks after dose one and 2-4 weeks after dose two, *n* = 144, 144, and 115, respectively. C–D) Comparison of anti-spike and anti-RBD IgG levels between residents (*n* = 116) and staff (*n* = 113) stratified by vaccine type at 2-4 weeks after dose two. Serum/plasma and DBS samples were diluted 4-fold to 0.0156 μL and 0.625 μL, respectively. The Kruskal-Wallis test and Dunn’s post test was used to determine significance. *** P < 0.001, * P < 0.05 after Bonferroni correction. Please note that the BNT162b2 post-dose 2 staff ELISA data were also reported in [33, 34].

To determine whether the vaccine type effect was also present in a younger (and presumably healthier) population, we obtained serum from LTCH staff, caregivers, and hospital-based healthcare workers after two doses of mRNA vaccine (*n* = 113; median age = 47). This population had higher humoral spike and RBD responses to both vaccines than the residents (*n* = 116; median age = 89); however, mRNA-1273 still yielded a stronger response than BNT162b2 (Figure 2C, D). Signal saturation was detected at both sample dilutions tested (Supplemental Figure 1C, D), further emphasizing the expected stronger humoral response in this group.

### mRNA-1273 induces higher neutralization of wild-type (D614G) than BNT162b2 in both LTCH residents and staff

Using a previously optimized [25] neutralization system from ref. [24] and a cohort of staff and resident samples taken 2–4 weeks post-dose 2, we monitored the neutralization of a wild-type spike-pseudotyped lentivirus (see Supplemental Figure 2 for example response curves). In both the staff and resident cohorts, the mRNA-1273 vaccine led to higher neutralization titers (2.95-fold and 3.90-fold higher than the BNT162b2 vaccine, respectively). Additionally, staff titers were higher than resident titers with both the mRNA-1273 (4.79-fold) and BNT162b2 (6.31-fold) vaccines (Figure 3A, Supplemental Figure 2). In multivariable analysis, older age and immunosuppression were associated with lower neutralization titers (Table 2), but did not interact with or modify the effect of vaccine type.

**Figure 3.**
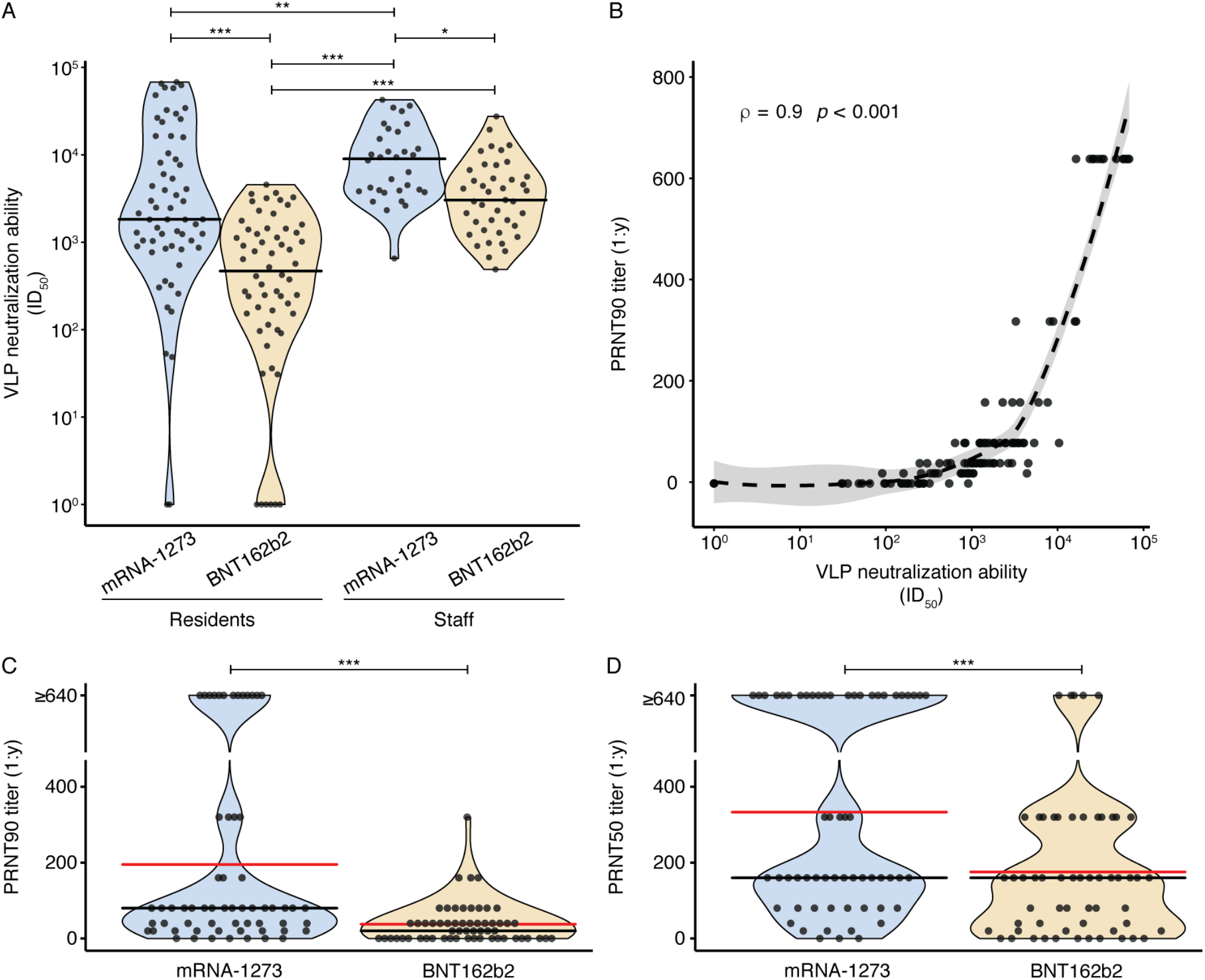
Neutralization of entry of wild-type D614G typed particles as defined in a S-pseudotyped lentiviral system. A) Neutralization ability in residents (*n* = 119) and staff (*n* = 78) stratified by vaccine type (mRNA-1273, Moderna; BNT162b2, Pfizer) 2–4 weeks post-dose two, as assessed by the VLP assay using lentiviral particles pseudotyped with wild-type spike. Resulting ID50 (50% neutralization titer) values are shown. The Kruskal-Wallis test and Dunn’s post test were used to determine significance. *** P < 0.001, ** P < 0.01, and * P < 0.05 after Bonferroni correction. B) Correlation between the ID50 values and PRNT90 titers of residents tested with both assays (*n* = 119). The dashed line indicates the locally weighted smoothing (LOESS) regression line, and grey shading indicates the 95% confidence interval. The Spearman’s rho coefficient and *P*-value are indicated. Note that all points indicating a PRNT90 titer of 1:640 are ≥1:640. C-D) PRNT90 and PRNT50 titers of resident samples stratified by vaccine type. Solid black lines indicate the median values for each violin, and solid red lines indicate the mean values for each violin. Mann-Whitney U test for significance was performed. ***P<0.001.

The lentiviral neutralization results were highly correlated (ρ = 0.9, *p* < 0.001) with those of a PRNT90 assay performed with wild-type SARS-CoV-2 on the same LTCH resident samples (Figure 3B). The mRNA-1273 vaccine induced higher neutralization titers than BNT162b2, and its median PRNT90 titer was higher (1:80) than that of BNT162b2 (1:20) (Figure 3C). More residents exhibited ≥1:640 PRNT50 titers with mRNA-1273 compared to BNT162b2; however, the median values were the same (1:160) (Figure 3D). In PRNT50 and PRNT90 neutralization assays, older age and immunosuppression were again associated with reduced responses to vaccination (data not shown). Notably, 3 of 14 residents with no detectable titers in the PRNT50 assay (including the one non-responder who received the mRNA-1273 vaccine) were immunosuppressed (P=0.01 compared to 2 of 112 other residents tested). Similarly, 4 of 33 without detectable PRNT90 titers were immunosuppressed (P=0.02 compared to 1 of 94 responders; the only immunosuppressed resident with detectable PRNT90 titers had previous had PCR-confirmed COVID-19).

### Several LTCH residents are unable to neutralize P.1 and B.1.351

We next used the same sera to test lentiviral virus-like particles (VLPs) derived from the B.1.1.7, B.1.351, P.1 and B.1.617.2 VOCs [26]. Residents who received BNT162b2 had fold-change reductions in neutralization of 1.67 (B.1.1.7), 2.88 (P.1), 4.25 (B.1.617.2) and 7.99 (B.1.351) compared to against VLPs derived from the wild-type strain, consistent with what has been previously reported [27, 28] (Figure 4; Supplemental Figures 2-4). Likewise, staff who received BNT162b2 showed a similar trend in neutralization reduction (1.88-, 2.55-, 2.91 and 4.80-fold for B.1.1.7, P.1, B.1.617.2 and B.1.351, respectively). Similar changes were observed for staff and residents vaccinated with mRNA-1273 (residents = 1.32, 2.17, 2.79 and 5.28-fold for B.1.1.7, P.1, B.1.617.2 and B.1.351, respectively; staff = 2.35, 3.30, 4.69 and 6.02).

**Figure 4.**
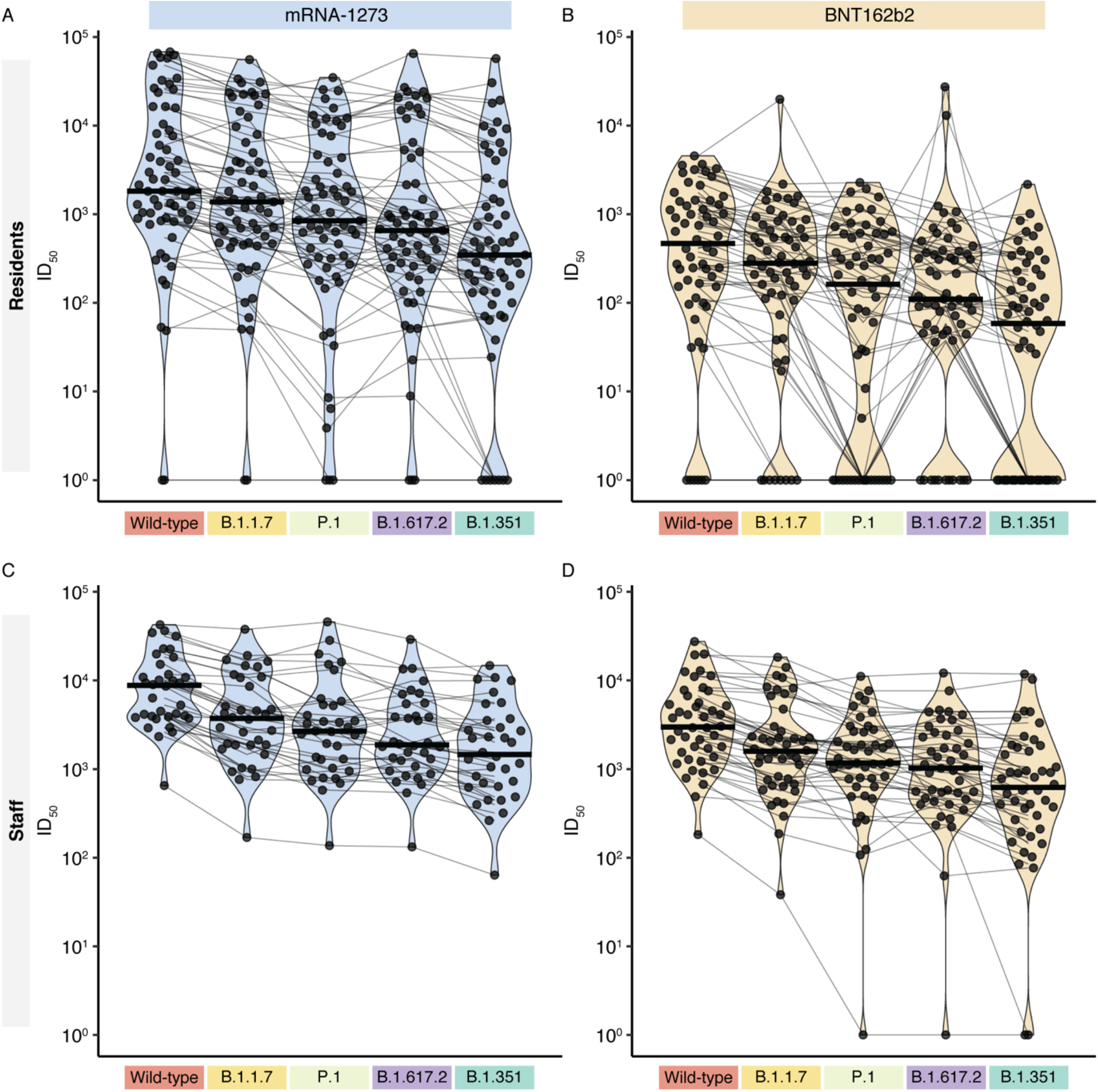
VOC lentiviral spike pseudotyped virus assay results by vaccine type and sample cohort. Violin plots show the distributions of ID_50_ values of samples taken from LTCH residents (top row, A-B) and staff (bottom row, C-D) 2– 4 weeks after dose two. Distributions are stratified by vaccine type (mRNA-1273, Moderna; BNT162b2, Pfizer; columns) and cohort [LTCH residents (*n* = 119) and staff (*n* = 78), rows]. Points represent individual VLP assay results, and points from identical samples are connected with thin grey lines. Thick black lines indicate the median values of each distribution.

Notably, multiple residents displayed no detectable neutralization. This was observed across all VLPs and in both vaccine cohorts, but was particularly pronounced with the B.1.351 and P.1 VOCs in the BNT162b2 cohort. In this cohort, 37.9% of the individuals (*n* = 22/58) were unable to neutralize B.1.351, compared to 11.5% (*n* = 7/61) of the mRNA-1273 cohort. Only 4.55 % (*n* = 2/44) of the staff BNT162b2 group did not respond, and the entire staff cohort receiving the mRNA-1273 vaccine displayed neutralizing responses to B.1.351. Similarly, 29.3% of residents who received the BNT162b2 vaccine had no response to P.1, compared to the 4.9% of residents administered the mRNA-1273 vaccine who did not elicit a response. All staff who received either vaccine showed detectable neutralization to P.1 except for one participant who received the BNT162b2 vaccine.

In summary, consistent with the differences in total antibodies induced by the mRNA-1273 and BNT162b2 vaccines in fully vaccinated residents and staff members, the mRNA-1273 vaccine induced stronger neutralization of all VOCs studied. In agreement with the literature, the wild-type strain was more readily neutralized than B.1.1.7, P.1, B.1.617.2 and B.1.351, and a large number of BNT162b2-vaccinated residents were unable to detectably neutralize P.1 and B.1.351.

### Assay correlations and establishment of thresholds for VOC neutralization

Participants with lower neutralization titers against the wild-type strain tended to also have lower—and in some instances no—neutralization of VOCs (Figure 4, connecting lines). Consistent with this, there were strong correlations (based on Spearman rho coefficients) between the wild-type strain and the VOCs, as well as between different VOCs (Figure 5, all ρ ≥ 0.89, *p* < 0.001). This high correlation enabled us to define thresholds for the detectable neutralization of a given VOC, if the neutralization ability against a different strain was known. For example, looking at the neutralization of the most immune evasive B.1.351 VLP, we determined a minimum ID_50_ level with each of the other VLPs that would be indicative of any B.1.351 neutralization (Figure 5, bottom row). This largely corresponded to the relative evasion of the different VLPs, with log_10_ID_50_ thresholds of 2.96 (ID_50_ = 906), 2.76 (ID_50_ = 582), and 2.57 (ID_50_ = 369) for wild-type, B.1.1.7, and P.1, respectively. Above these thresholds, at least some neutralization activity was detected for B.1.351, which largely correlated with the titers for neutralization of the other VOC-typed VLPs. This indicates that if the neutralization titer of only one lentivirus is known and passes a minimum level, the neutralization of other VOCs can be inferred reasonably well.

**Figure 5.**
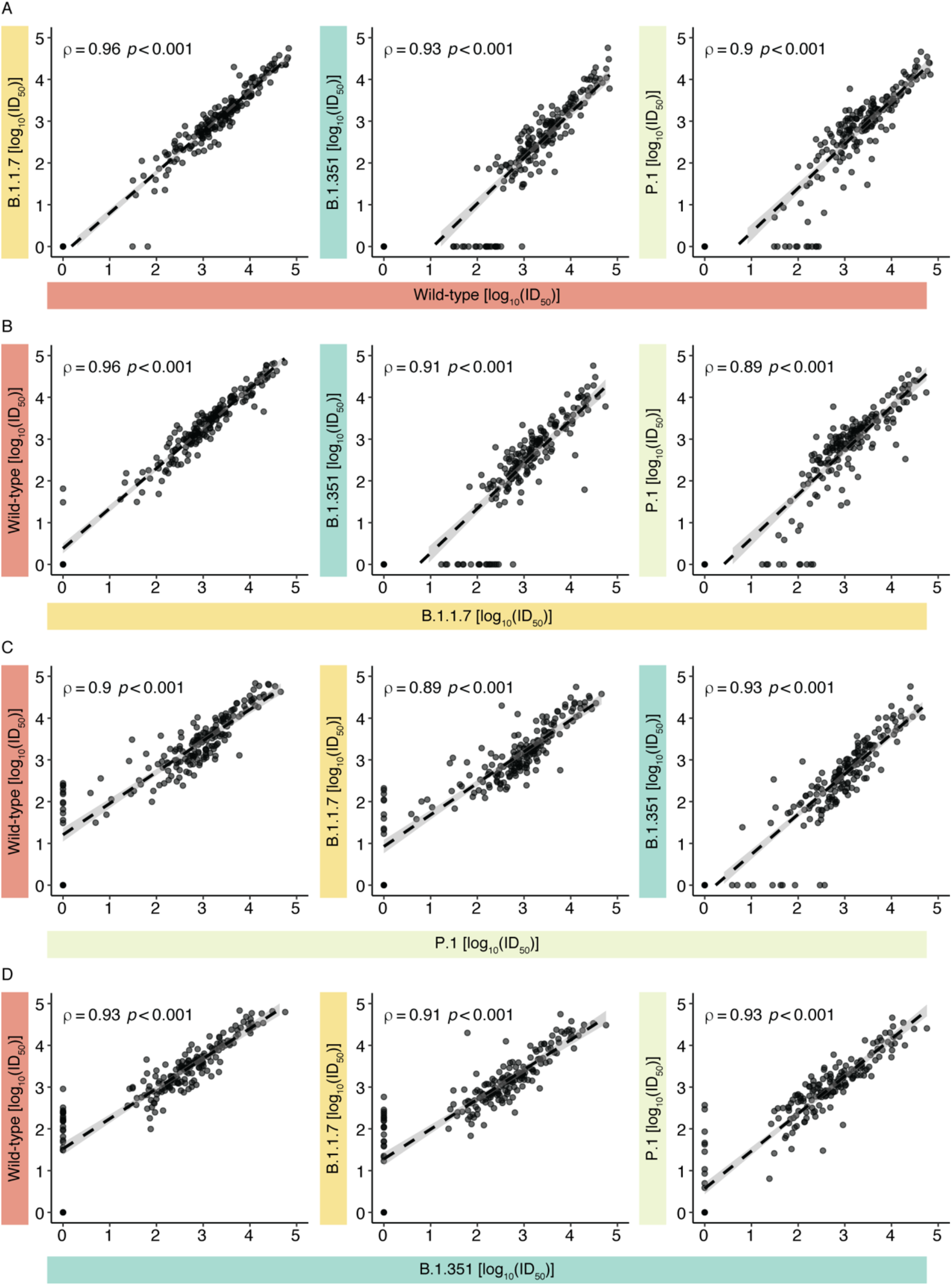
Correlations between VOC neutralization titers as determined by the VLP neutralization assay. Neutralization results (*n* =197) are expressed as log_10_ID_50_ values. The dashed line indicates the regression line, and grey shading indicates the 95% confidence interval. Spearman’s rho coefficients and *P*-values are indicated.

We next investigated whether anti-spike and/or anti-RBD IgG levels could also be used to infer neutralization potential against VOCs, since it is generally much easier to measure these values on a large scale [23]. Spearman’s correlation coefficients indicated strong correlations between VLP neutralization and anti-RBD/spike IgG levels (Figure 6), as well as between overall anti-spike and anti-RBD levels (Supplemental Figure 5). With the wild-type- and B.1.1.7-typed particles, participants who were seropositive for anti-spike or anti-RBD antibodies generally showed neutralizing capabilities with the VLP assay that correlated well with their relative IgG levels. However, the seropositivity threshold was less predictive when looking at neutralization titers for the B.1.351 and P.1 variants. Notably, there was a subgroup of participants whose anti-IgG and anti-RBD levels were well above the threshold for seropositivity (and even the median of convalescents for some) yet could not neutralize VOC-typed VLPs. Thus, accurate prediction of VOC neutralization requires a better definition of the thresholds for each VOC. We assessed threshold values for anti-RBD and anti-spike IgG levels by calculating receiver operator characteristic (ROC) curves for each antigen across the VOCs (Supplemental Figure 6). On each ROC curve, we determined the optimal cutoff by calculating the top-left most point of each curve. For the wild-type and B.1.1.7 strains, the RBD and spike had comparable areas under the curve (AUCs) and similar specificities and sensitivities (Supplemental Table 1). For spike, a threshold of 0.91 yielded a specificity of 1.00 and sensitivity of 0.92 for the wild-type strain with near identical parameters with B.1.1.7, and this threshold was much higher than the seropositivity threshold of 0.19. Optimal thresholds for P.1 and B.1.351 were further increased to 1.37 and 1.47, respectively, and both specificities and sensitivities were reduced compared to the wild-type strain. Notably, these thresholds are similar to the median level of 1.38 in a cohort of convalescent patients. Similarly, the anti-RBD thresholds increased with each VOC. A threshold level of 0.14 was optimal to classify neutralization of the wild-type strain, which was below the threshold of 0.18 required for seropositivity. However, this threshold increased to 1.01 with B.1.351 (which is still below the threshold of 1.25 in convalescent patients).

**Figure 6.**
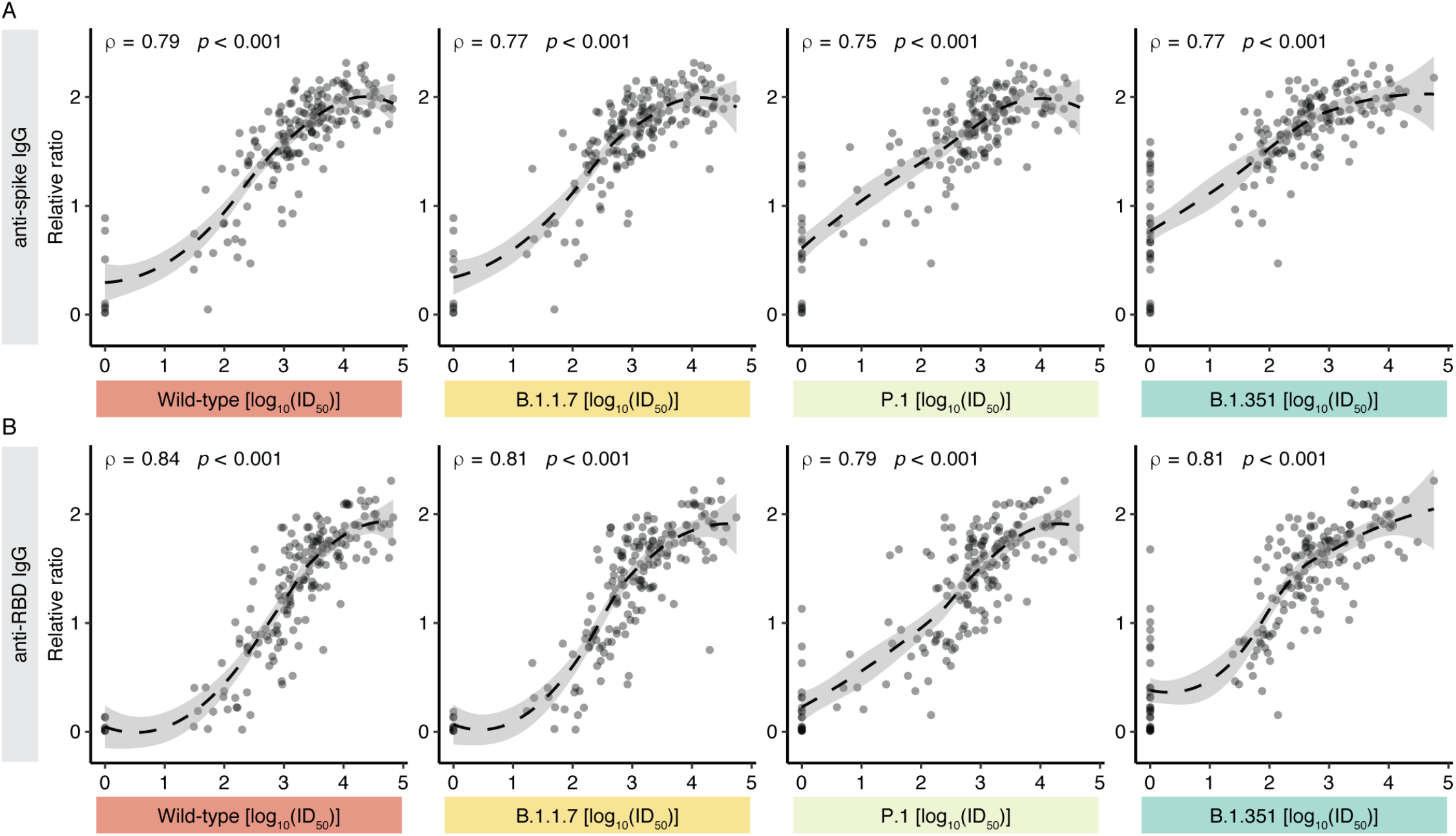
Correlations between anti-spike/RBD IgG responses and neutralization as determined by the VLP neutralization assay. IgG responses (*n* = 192) are expressed as relative ratios and neutralization results are expressed as log_10_ID_50_ values. The dashed line indicates the regression line fit using LOESS, and grey shading indicates the 95% confidence interval. Spearman’s rho coefficients and P values are indicated.

In summary, despite the high correlation between the VOC neutralization responses and IgG levels, it was difficult to use a single metric to predict whether a participant might be able to neutralize a VOC. However, the increasingly higher thresholds derived from the precision-recall curves suggest that higher levels of anti-spike and anti-RBD antibodies are required for protection against VOCs.

## Discussion

This work joins other studies [8-11] (often with smaller cohorts) in demonstrating that frail older adults do not mount as strong an immune response to COVID-19 vaccines as younger and healthier individuals. Our study also corroborates findings in other cohorts that mRNA-1273 elicits a stronger humoral response (in terms of total and neutralizing antibodies) than BNT162b2. In the LTCH resident cohort, vaccination with mRNA-1273 yielded fewer individuals that failed to mount a detectable neutralization response, and this was also corroborated in a recent study in Ontario LTCH [29]. The reasons for the difference in the humoral responses of the vaccines have not been explored in depth here. One possibility is that the mRNA dose may influence the strength of the humoral immune response to spike – 100 µg of mRNA is administered with the mRNA-1273 vaccine which is 3.3-fold higher than the BNT162b2 vaccine.

The differences between the two vaccines were even more evident when the neutralization of VOCs was considered (rather than that of the wild-type strain, against which the vaccines were designed). For example, 37.9% of BNT162b2-vaccinated LTCH residents displayed no detectable neutralization of the B.1.351 strain, compared to 11.5% for those who received mRNA-1273. This is a sobering finding in light of the remarkable efficiency in these two vaccines in building high neutralization titers in clinical trials compared to other vaccines used around the world [16].

Despite the high correlation between neutralization and vaccine efficacy, neutralizing antibodies represent only one facet of the immune response to vaccination [30]. In healthy individuals, the first dose of vaccine does induce good protection against severe disease (although weaker than the two-dose regimen) and symptomatic infection associated with different variants [31, 32], despite lower neutralization levels than elicited by the full 2-doses. Binding (non-neutralizing) antibodies can promote Fc-mediated effector functions that contribute to vaccine efficacy, and these are likely to be less affected by VOCs, as are T cell-mediated responses [27]. While we have not directly examined the T cell response or Fc-receptor functions here, binding antibodies were monitored, which also suggested weaker responses in BNT162b2 recipients than in mRNA-1273 recipients at all time points, and a clear difference in the responses of LTCH residents and staff 2–4 weeks after dose 2.

There are some other limitations to this study: we have only analyzed the response to COVID-19 the mRNA vaccines, with both doses administered on the schedule recommended by the vaccine manufacturers. It is noteworthy that in Ontario, LTCH residents were prioritized for vaccination with mRNA vaccines administered according to the product monograph. However, LTCH residents in some other provinces of Canada, as well as many older adults and individuals with immune system deficiencies have been vaccinated across the country with two doses of mRNA vaccines spaced further apart in order to accommodate vaccine supply issues. Another segment of the population has been vaccinated with the ChAdOx1-S vaccine (from AstraZeneca or COVISHIELD), which yields lower neutralizing antibodies [16]. Importantly, heterologous dose regimens (particularly for those who received ChAdOx1-S vaccine as a first dose) have been widely used in Canada, but their effect on the humoral response in older adults is still not known. Furthermore, we have only looked at the response 2–4 weeks after dose 2, and it will be critical to profile the decline in antibody production in this priority population. Lastly, it is possible that as the virus continues to evolve, new variants may further evade or overwhelm the blunted humoral response generated by vaccination in these older and frail adults.

In conclusion, our results raise further concern that even the most effective current vaccines provide only partial protection to individuals with known vulnerabilities in the face of a pandemic now largely driven by VOCs, particularly B.1.617.2. We suggest that longitudinal monitoring of the antibody response as well as other components of the immune response to SARS-CoV-2 be performed, and that vaccination strategies for individuals with known vulnerabilities should consider which vaccine and at what dose and frequency is administered for optimal immunity in priority populations.

## Methods

### Participant recruitment

Residents, their essential caregivers, and staff at four LTCHs located in south-central Ontario who intended to get a COVID-19 vaccine were eligible to participate in the study. Study staff approached LTCH administrators in the greater Toronto area to identify interest. Interested administrators were asked to notify residents, their substitute decision makers, and staff about the study, and to offer them the choice of being contacted by study staff or opting out of all study contact. The administrator shared contact information for those who did not opt out with study staff. The study team then contacted staff and residents/substitute decision makers for consent to participate. Participants were asked to consent to blood draws via venipuncture and/or dried blood spots via fingerprick. Participants could decline any individual blood draw at the time of sampling; specimens were also not obtained from residents who were hospitalized for any reason at the time of sampling. Baseline demographic, clinical, and vaccine data, including information on SARS-CoV-2 infection prior to the study, were obtained from participants either through telephone interview (staff, caregivers) or chart review (residents). During study visits and follow up calls, study participants’ SARS-CoV-2 infection status was continuously ascertained via chart review and/or reports from LTCH staff and caregivers.

#### Sample collection

For baseline samples and samples collected 3–4 weeks post-dose 1 (and for the samples post dose 2 in participants who declined venipuncture or from whom venipuncture could not be obtained), DBS samples were collected on Whatman 903 Protein Saver cards (GE Healthcare, Boston, MA, USA). For participants who agreed to venipuncture, a 10 ml serum sample was obtained 14–28 days post-dose 2 (in rare cases of scheduling conflicts, samples are accepted up to 42 days post-dose 2). Samples were aliquoted and frozen at -80°C.

Please note that the BNT162b2 post-dose 2 staff ELISA data were also reported in [33, 34].

### Automated ELISAs

We previously reported the development of ELISA-based assays in 384-well chemiluminescent format to assess the persistence of antibody responses in infected individuals [23]. We have since modified the assays to use a more stable source of antigens and IgG calibration and detection reagents, all generated at the National Research Council of Canada (Colwill et al., in preparation). The reformulated assays were validated as in ref. [23] by ROC curve analysis using both pre-COVID era negative controls and sera/plasma from qPCR-confirmed convalescents, and the cutoffs for seropositivity for individual assays were defined as those that pass both a 1% false positive rate (FPR) threshold and exhibit ≥3 standard deviations (SDs) from the log means of the negative controls (relative ratio to the synthetic standard; see below) performed over 4 months on the same system. Because serum/plasma and DBS samples have different background luminescence interference, their thresholds needed to be calculated separately, and are indicated in the figures (for DBS samples, this value was first corrected as detailed below). For DBS samples, we used a 1% FPR threshold determined by duplicate ROC analysis of 90 negative and 97 positive DBS samples (a generous gift of the National Microbiology Laboratory of Canada). The median of the convalescent sera/plasma samples (Colwill et al., in preparation) is indicated in the figures as a reference point.

#### Test sample preparation

DBS samples were punched (2 × 3mm) using a BSD600 Ascent semi-automated puncher (BSD Robotics, Brisbane, QLD, Australia) and antibodies were eluted in PBS plus 0.1% Tween-20 (PBST) supplemented with 1% Triton X-100 for a minimum of 4 hours with shaking at 150 rpm. Eluates were centrifuged at 1,000 × *g* for 30 seconds, then diluted 1:4 in 1.3% Blocker BLOTTO (Thermo Fisher Scientific, Waltham, MA, USA) to a final concentration of 1% Blocker BLOTTO in PBST. Samples submitted as frozen serum or plasma were treated with Triton X-100 to a final concentration of 1% for 1 hour prior to use.

#### Standard curves

We used recombinant antibodies able to detect both spike and its RBD (IgG: VHH72hFc1X7; National Research Council of Canada). For NP, we used human anti-nucleocapsid IgG (#A02039, clone HC2003, GenScript, Piscataway, NJ, USA). In all cases, a 8-point 4-fold dilution series was added to each plate (see below for volumes and dilution buffers).

#### Positive and negative controls

In addition to the standard curves listed above, a negative control containing purified IgG from human serum (final concentration 1 µg/mL; #I4506, Millipore-Sigma, Oakville, ON, Canada) and pools of positive (from convalescent patients with high IgG levels to all three antigens) and negative sera (from 3–4 patients) were added to each plate as 2-point 4-fold dilutions for quality control and to enable cross-plate comparisons.

All washes were performed 4 times in 100 μL PBST, using liquid dispensers (Biomek NX^P^ Automated Liquid Handling Workstation, Beckman Coulter, Indianapolis, IN, USA; Multidrop Combi Reagent Dispenser, Thermo Fisher Scientific) and a 405 TS/LS LHC2 Microplate Washer (BioTek Instruments, Winooski, VT, USA) on an F7 Robot System (Thermo Fisher Scientific) at the Network Biology Collaborative Centre (nbcc.lunenfeld.ca). Antigens were diluted in 10 μL PBS and dispensed into Greiner Bio-One 384-Well F-Bottom Polystyrene Microplates (#781074, Greiner Bio-One, Monroe, NC, USA) at final amounts of 50 ng spike, 25 ng RBD, and 7 ng NP per well. The plates were centrifuged at 233 × *g* for 1 minute to ensure even coating, incubated overnight at 4°C, and washed. Wells were blocked with 80 µL of 5% Blocker BLOTTO in TBS (Thermo Fisher Scientific, #37530) for 1 hour and then washed. Samples and controls were diluted in 1% Blocker BLOTTO in PBST, and 10 µL was added to each well from 96- or 384-well source plates. Plates were incubated for 2 hours at room temperature and wells were washed with PBST. Secondary antibodies (described below) were diluted as indicated in 1% Blocker BLOTTO in PBST and 10 µL was added to each well. After incubation for 1–2 hours at room temperature, the wells were washed and 10 µL of SuperSignal ELISA Pico Chemiluminescent Substrate (Thermo Fisher Scientific, #37069, diluted 1:4 in Milli-Q dH_2_0) was dispensed to each well and mixed at 900 rpm for 10 seconds. After a 5–8 minute incubation, plates were read on an EnVision 2105 Multimode Plate Reader (Perkin Elmer, Woodbridge, ON, Canada) at 100 ms/well using an ultra-sensitive luminescence detector.

#### Quality control assessment

For each automated test, the raw values and relative ratios of each control are compared to controls from prior tests to confirm that the values are similar. Our criteria were that the raw values of the reference points and positive pool controls compared to the blanks must be within 90% of previous runs and the reference points for each antigen compared to each other must be within 90% of previous runs. The log_10_ sample density distribution (raw values and relative ratios) was compared to prior runs to confirm that the distribution was within range.

#### Expression of the results

Raw results (in luminescence units) were expressed as ratios to a blank-adjusted point in the linear range of the relevant recombinant antibody standard reference curve. All values are calculated using automated scripts, prior to consideration of the meta-data; the investigators performing the measurements were blinded to the sample description.

#### DBS/serum data transformation

Fifteen participants had DBS and serum samples taken at the same time, and each sample was processed at two dilutions. To compare DBS and plasma/serum samples, we applied a normalization factor to account for the differences in sampling methods. Using the equation of the regression line, we converted results from samples processed using 2.5 µL DBS eluate to values reflected by samples processed using 0.0625 µL serum/plasma. To convert DBS eluate values to serum/plasma values, we used the equation y = 0.05 + 1.1x (R^2^ = 0.99, *p* = 4.4e-12) for the RBD and y = 0.044 + 1.1x (R^2^ = 0.98, *p* = 1.6e-10) for spike. Similarly, we converted values generated from 0.625 µL DBS eluate to 0.0156 µL serum/plasma. For this conversion, we used the equation y = 0.14 + 1.1x (R^2^ = 0.96, *p* = 5.5e-9) to convert between RBD levels and the equation y = 0.093 + 1.1x (R^2^ = 0.86, *p* = 4.9e-6) to convert between spike levels, where y is the relative ratio of DBS using 0.625 µL of the eluate and x is the relative ratio using 0.0156 µL of plasma/serum samples. DBS thresholds were adjusted accordingly. Data were cleaned, analyzed, and plotted using R (v. 4.1.0).

#### Using baseline ELISA results to support evidence of prior infection

Samples that passed the 3-SD threshold with ≥ 2 antigens at the baseline (pre-vaccination) point were considered to have evidence of previous infection.

### Spike-pseudotyped lentivirus neutralization assays

The generation of pseudotyped lentivirus particles and the pseudovirus neutralization assay were performed as described previously [25, 26], with some modifications. Pseudotyped lentivirus particles were generated from wild-type SARS-CoV-2 (defined as the Wuhan Hu-1 sequence harboring the commonly circulating D614G mutation) and the VOCs B.1.1.7, B.1.351, and P.1 spike protein constructs were previously described [26]. The B.1.617.2 sequence (derived from https://outbreak.info/compare-lineages?pango=B.1.617.2&gene=S&threshold=0.2) was codon-optimized for expression in human cells and synthesized (Twist Bioscience), transferred to the HDM lentiviral backbone, and fully sequenced. Each spike construct was co-transfected with packaging (psPAX2, Addgene, Watertown, MA, USA, #12260) and luciferase reporter constructs (pHAGE-CMV-Luc2-IRES-ZsGreen-W, kindly provided by Jesse Bloom) into HEK293TN cells (System Biosciences, Palo Alto, CA, USA, LV900A-1). Following 8 hours of transfection, cell supernatants were replaced with 3 mL of viral production medium containing 5% heat-inactivated FBS and 1% penicillin/streptomycin, and incubated at 37°C and 5% CO_2_ for 16 hours, then at 33°C and 5% CO_2_ for an additional 24 hours. Viral supernatants were harvested, clarified, and filtered through 0.45 µm filters prior to storage at -80°C. A viral titer assay was performed using HEK293-ACE2/TMPRSS2 cells, and a virus dilution resulting in > 1,000 relative luciferase units over the control was chosen (∼1:100 of virus stock; ∼15% infection of seeded cells). HEK293TN and HEK293-ACE2/TMPRSS2 cells were maintained at 85% confluency for no more than 25 passages.

For the neutralization assay, patient sera were first heat inactivated at 56°C for 30 minutes and diluted in DMEM (containing 10% heat inactivated FBS and 1% penicillin/streptomycin) at 1:20 (or 1:100 for samples that displayed higher neutralization activity; Supplemental Table 1), followed by 2.5-fold serial dilutions (8 dilutions total), and incubated with diluted virus at a 1:1 ratio for 1 hour at 37°C prior to addition to HEK293-ACE2/TMPRSS2 cells. Cells were incubated for an additional 48 hours, followed by lysis and detection of luminescence signals using the Bright-Glo Luciferase Assay System (Promega, Ottawa, ON, Canada, E2620) and the EnVision 2105 Multimode Plate Reader. Unless otherwise specified, 50% neutralization titer (ID_50_) values of patient sera were calculated in GraphPad Prism 9 (GraphPad Software, San Diego, CA, USA) using a nonlinear regression (log[inhibitor] versus normalized response – variable slope) algorithm. All measurements were generated blinded to the metadata (including vaccine type), though the staff versus resident designation was known to the experimentalists (naming convention).

### PRNT assays

SARS-CoV-2 (Canada/ON_ON-VIDO-01-2/2020, EPI_ISL_42517) stocks were titrated as previously reported [25] [35]. The SARS-CoV-2 PRNT assay was adapted from a previously described method for SARS-CoV-1 [36]. In a 96-well plate, serially diluted antibodies were mixed with diluted SARS-CoV-2 (100 PFU/100 μL), yielding final antibody dilutions of 1:20 to 1:640 and a final virus concentration of 50 PFU/100 μL. No neutralization, 50% neutralization, and 90% neutralization controls were prepared by diluting SARS-CoV-2 at 50, 25, and 5 PFU/100 μL, respectively. DMEM supplemented with 2% FBS and 1× penicillin was used as a no virus control. After 1 hour of incubation at 37°C and 5% CO_2_, 100 μL of each antibody-virus mixture was added in duplicate to 12-well plates containing Vero E6 cells at 95–100% confluence, and 100 μL of each control was added in triplicate to two sets of 12-well plates containing Vero E6 cells. All plates were incubated at 37°C and 5% CO_2_ for 1 hour of adsorption, followed by the addition of a liquid overlay containing equal volumes of 3% carboxymethylcellulose (CMC) and 2× Modified Eagle’s Medium (Temin’s modification). After a 3-day incubation, the liquid overlay was removed and cells were fixed with 10% neutral-buffered formalin. The monolayer in each well was stained with 0.5% crystal violet (w/v) and the average number of plaques was calculated at each dilution and compared with the average number of plaques in the 50% and 90% neutralization controls. The reciprocal of the highest dilution resulting in 50% and 90% reduction in plaques compared with controls were defined as the PRNT50 and PRNT90 endpoint titers, respectively. PRNT50 titers and PRNT90 titers ≥ 20 were considered positive for SARS-CoV-2 neutralizing antibodies, whereas titers <20 were considered negative.

### Analysis of factors associated with humoral response to COVID-19 vaccines

Data on clinical response and antibody titers were entered into excel, cleaned and analyzed in SAS version 0.4 for PC (SAS Institute, Cary, NC). Based on knowledge of response to vaccines in adult populations in general, and available data for response to COVID-19 vaccines in other populations, we *a priori* selected age, sex, Charlson co-morbidity index score, presence or absence of immunosuppression (defined as per [37]), presence or absence of renal disease and body mass index (BMI) as variables to be assessed. The relationship of age and Charlson scores to antibody titers appeared linear; few residents had Charlson scores >5 such that we assigned a maximum Charlson score of 5 for the analysis. Only 5 residents were underweight, and there was no obvious correlation with antibody titers; titers declined above 35 kg/m^2^, such that BMI was modelled as normal or underweight versus severely or morbidly obese. Bivariate comparisons used proc corr for continuous variables, and Wilcoxon rank sum scores for categorical variables. Multivariable analysis was performed using proc glm and stepwise backwards elimination of variables. Residuals were examined to ensure homoscedasticity, and the absence of multicollinearity was confirmed with variance inflation factors being <5 for all predictors. P values of <0.05 were considered statistically significant.

## Data Availability

Additional data is available from the authors

## Study approval

REB approval was received from the Sinai Health System (20-0339-E). Informed consent was obtained from LTHC staff, essential caregivers and residents or their substitute decision makers.

## Author contributions

KTA performed data analysis, wrote software code, generated figures, and edited the manuscript. QH generated and analyzed the lentivirus neutralization data, and generated supplemental figures. MM performed data analysis and modelling, and contributed to metadata curation. These three authors shared the first position because their important and complementary contributions made the study possible; the order of the authors was defined by relative roles in assembling the manuscript. RS developed the neutralization assay and helped perform it. KM and AR performed the PRNT assays. KG, LG, SB, NH, AT, KQL, JGT, and CF participated in study design, REB development, participant recruitment, and data collection. AL and AP processed serum and plasma samples, and transferred them to the analytical laboratory. BR and MFZ prepared all samples for ELISA and tracked all samples within the analytical laboratory. MI and JW performed automated ELISAs. WRH developed the B.1.617.2 lentiviral constructs. YA and EM provided VOC lentiviral constructs. AP developed software for automated ELISA analysis. YD provided the proteins (antigens and antibodies) used in the automated ELISAs. GC, SSM, MO, and JLG contributed to the collection of LTCH staff samples. KC performed data analysis and supervised the ELISA data acquisition. SES supervised the project, organized sample collection, and acquired funding. HW supervised the PRNT assays and analyzed data. AJM supervised the project, organized sample collection, performed data analysis, wrote the manuscript, and acquired funding. ACG supervised the project, developed the analytical assays, performed data analysis, wrote the manuscript, and acquired funding.

## Acknowledgements

We are grateful to Kate Crawford and Jesse Bloom for the original lentiviral S-neutralization system and to High-Fidelity Science Communications for editing support.

This work was supported by a COVID-19 Immunity Task Force (CITF) award to Sharon E. Straus, Allison J. McGeer, Jennifer L. Gommerman, Mario Ostrowski and Anne-Claude Gingras (Wellness Hub). This work was also supported by funding from the Canadian Institutes of Health Research and Public Health Agency of Canada (through the CITF) to Anne-Claude Gingras, Allison J. McGeer and Mario Ostrowski (VOC neutralization in LTC settings). We also acknowledge support from Ontario Together and funding from the Canadian Institutes of Health Research (#VR1-172711 and #439999). Funding for initial assay development in the Gingras lab was provided through generous donations from the Royal Bank of Canada and the Krembil Foundation (via the Sinai Health Foundation). The robotics equipment used is housed in the Network Biology Collaborative Centre at the Lunenfeld-Tanenbaum Research Institute, a facility supported by the Canada Foundation for Innovation, the Ontario Government, and Genome Canada and Ontario Genomics (OGI-139). The Pandemic Response Challenge Program of the NRC supported the design, production, purification and analysis of the antigens and antibodies used in the automated ELISA assays. The B.1.617.2 (Delta) lentiviral assay was developed through funds from the Coronavirus Variants Rapid Response Network (CoVaRR-Net). Anne-Claude Gingras, Sharon Straus and Jennifer L. Gommerman are Canada Research Chairs (Tier 1) in Functional Proteomics; Knowledge Translation and Quality of Care; and Tissue Specific Immunity, respectively. Anne-Claude Gingras and Jennifer L. Gommerman are pillar leads for CoVaRR-Net.

---

**Supplemental Figure 1.**
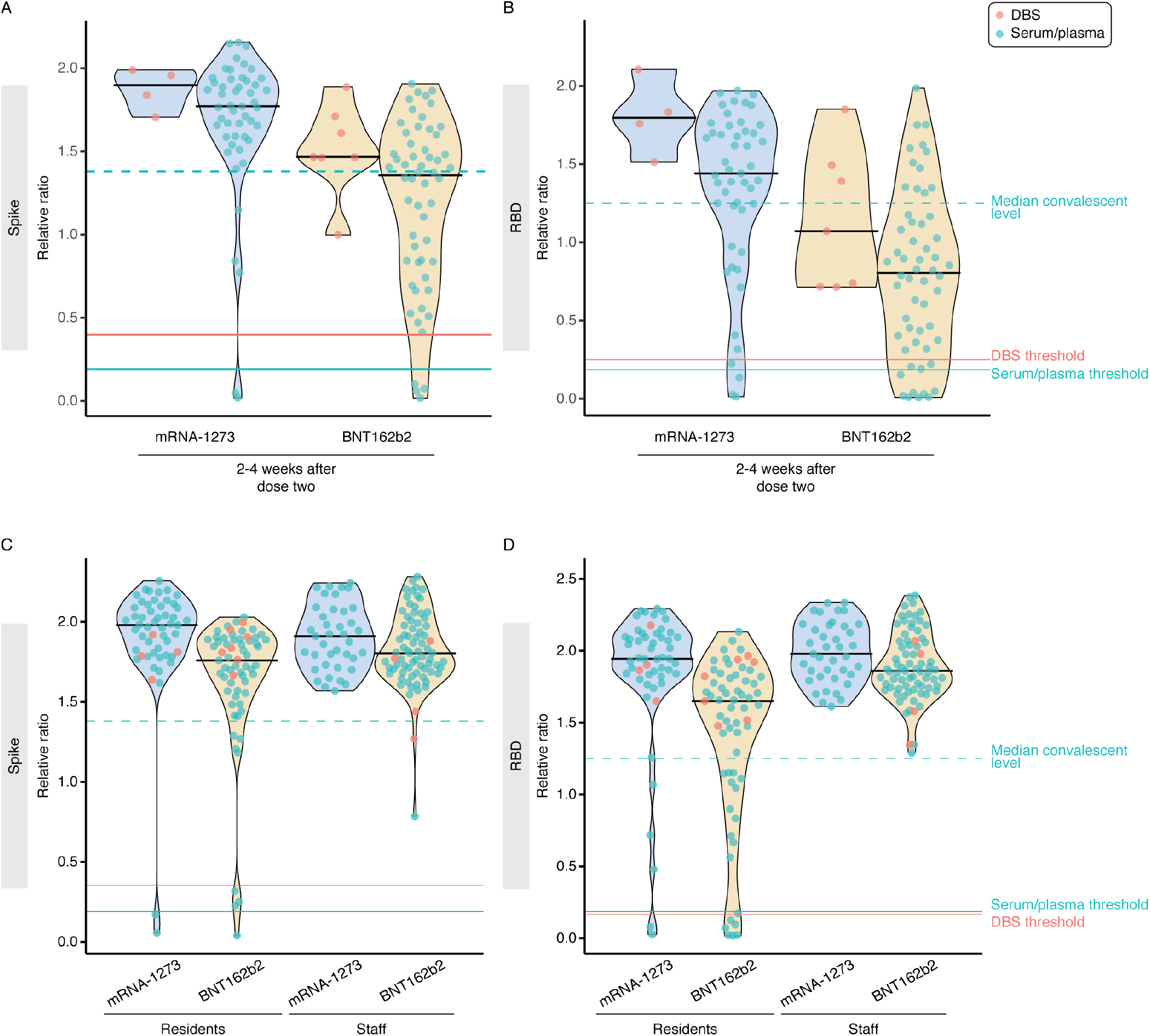
Contributions to response variability in vaccinated individuals. A-B) Levels of serum anti-spike and anti-RBD IgG levels stratified by vaccine type at a single time point (2-4 weeks after dose two) in not previously infected individuals. Turquoise and salmon points are results from serum/plasma (0.0156 μL sample used) and ratio-corrected DBS eluate (0.625 µl sample used), respectively. C-D) Comparison of anti-spike and anti-RBD IgG levels between residents and staff stratified by vaccine type at 2-4 weeks after dose two. Serum/plasma and DBS samples were processed at 0.0625 μL and 2.5 μL, respectively. This figure is related to Figure 2.

**Supplemental Figure 2.**
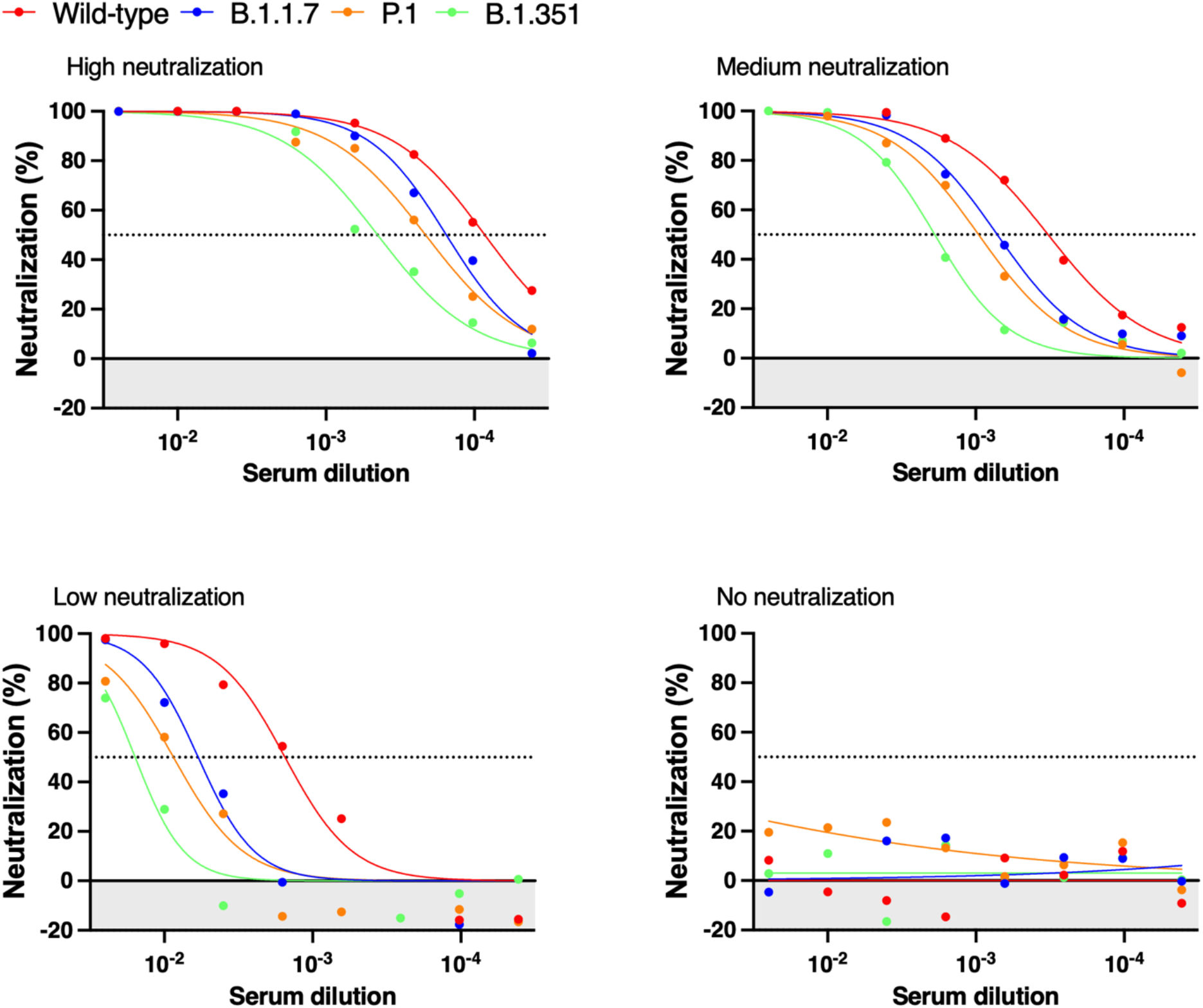
Representative cross-variant neutralization curves in high, medium, low, and non-neutralized individual samples. Normalized percentage neutralization values from individual LTCH staff and residents 2–4 weeks post-dose two and plotted against the logarithm of the dilution factors. Each plot represents one individual sample with high, medium, low, or no neutralization across the variants. Data points below 0 represent RLU values greater than the positive control, indicating no neutralization in sera samples (shaded area).

**Supplemental Figure 3.**
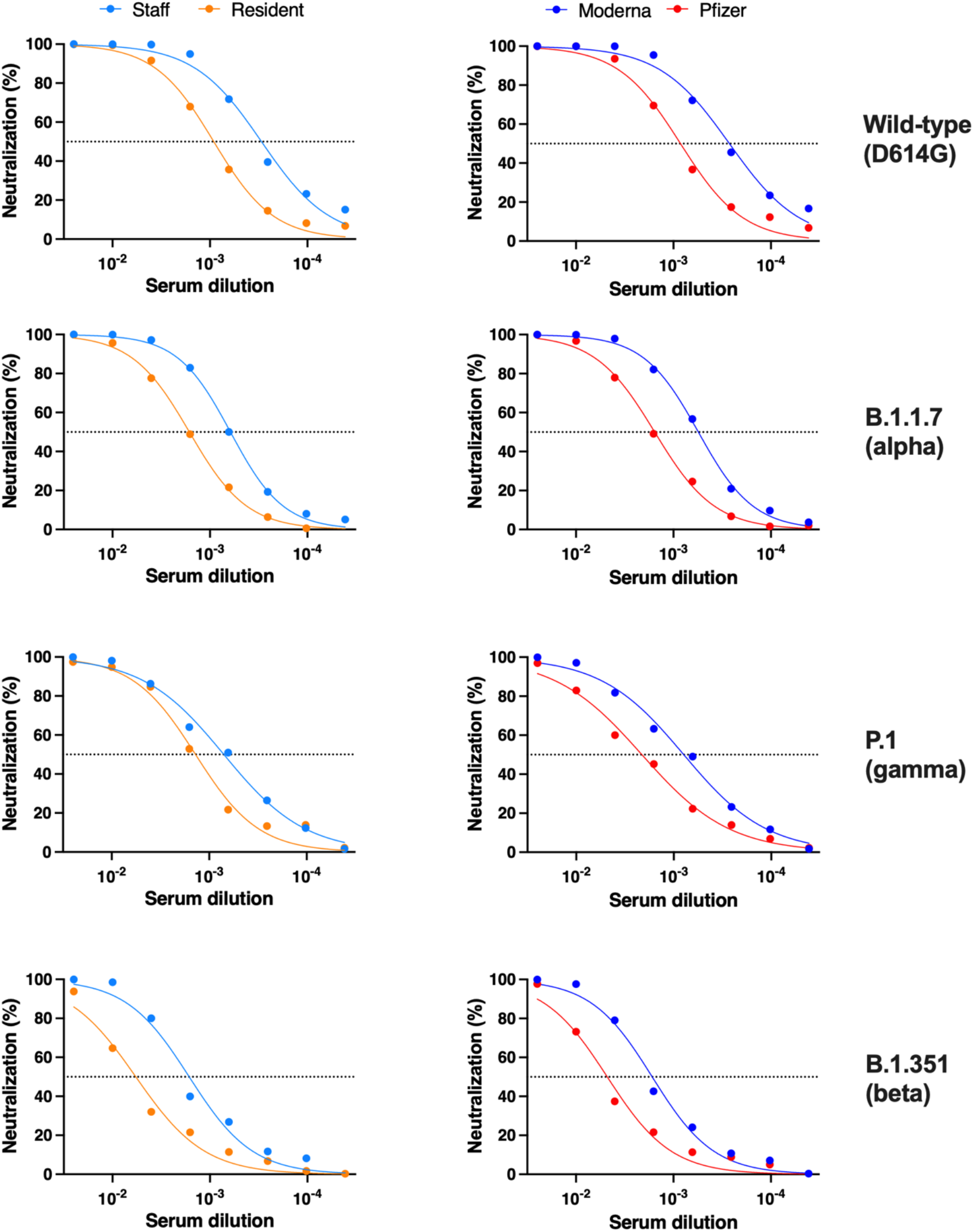
Detailed neutralization curves for VOCs. Left: Comparative pseudovirus neutralization curves between residents vs. staff. Right: Comparison between mRNA-1273 (Moderna) and BNT162b2 (Pfizer). Normalized percentage neutralization values were calculated, and the neutralization curves were plotted against the dilution factors against VOCs (wild-type, B.1.17, P1, and B1.351). Samples were from LTCH staff (*n* = 113), residents (*n* = 119), mRNA-1273-vaccinated individuals (*n* = 85), and Pfizer vaccinated individuals (*n* = 93). Data points represent the medians of each stratified group.

**Supplemental Figure 4.**
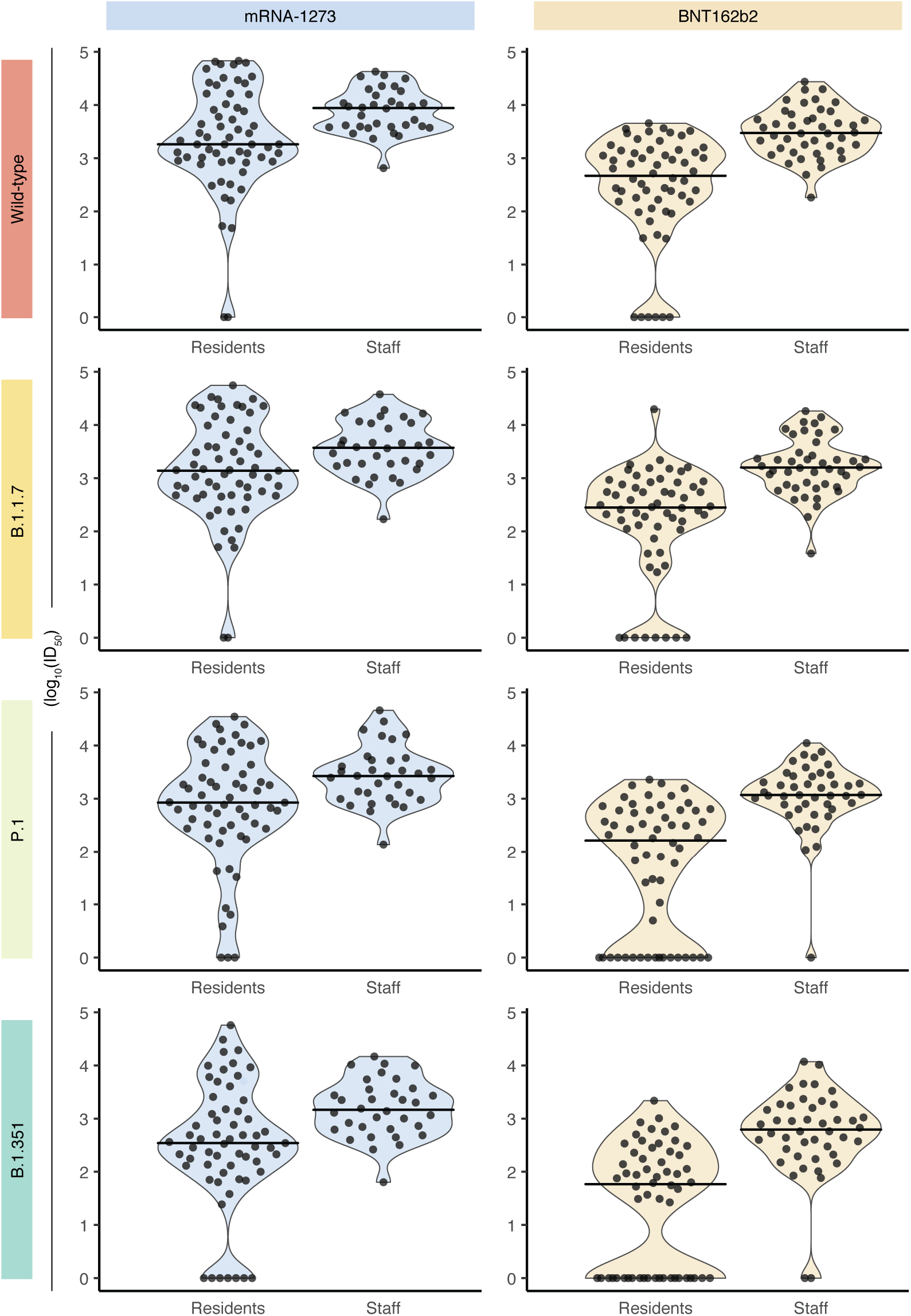
A comparison of lentiviral spike pseudotyped virus assay results between LTCH residents and staff stratified by vaccine type and variants of concern (VOC). Violin plots show the distribution of log_10_–transformed ID_50_ values of samples taken from LTCH residents (*n* = 119) and staff (*n* = 78) 2–4 weeks after receiving a second vaccine dose. Distributions are stratified across vaccine type [mRNA-1273 and BNT162b2, columns] and VOCs (Wild-type, B.1.17, P1 & B1.351, rows). Points represent individual VLP assay results, and lines indicate the median values of each distribution.

**Supplemental Figure 5.**
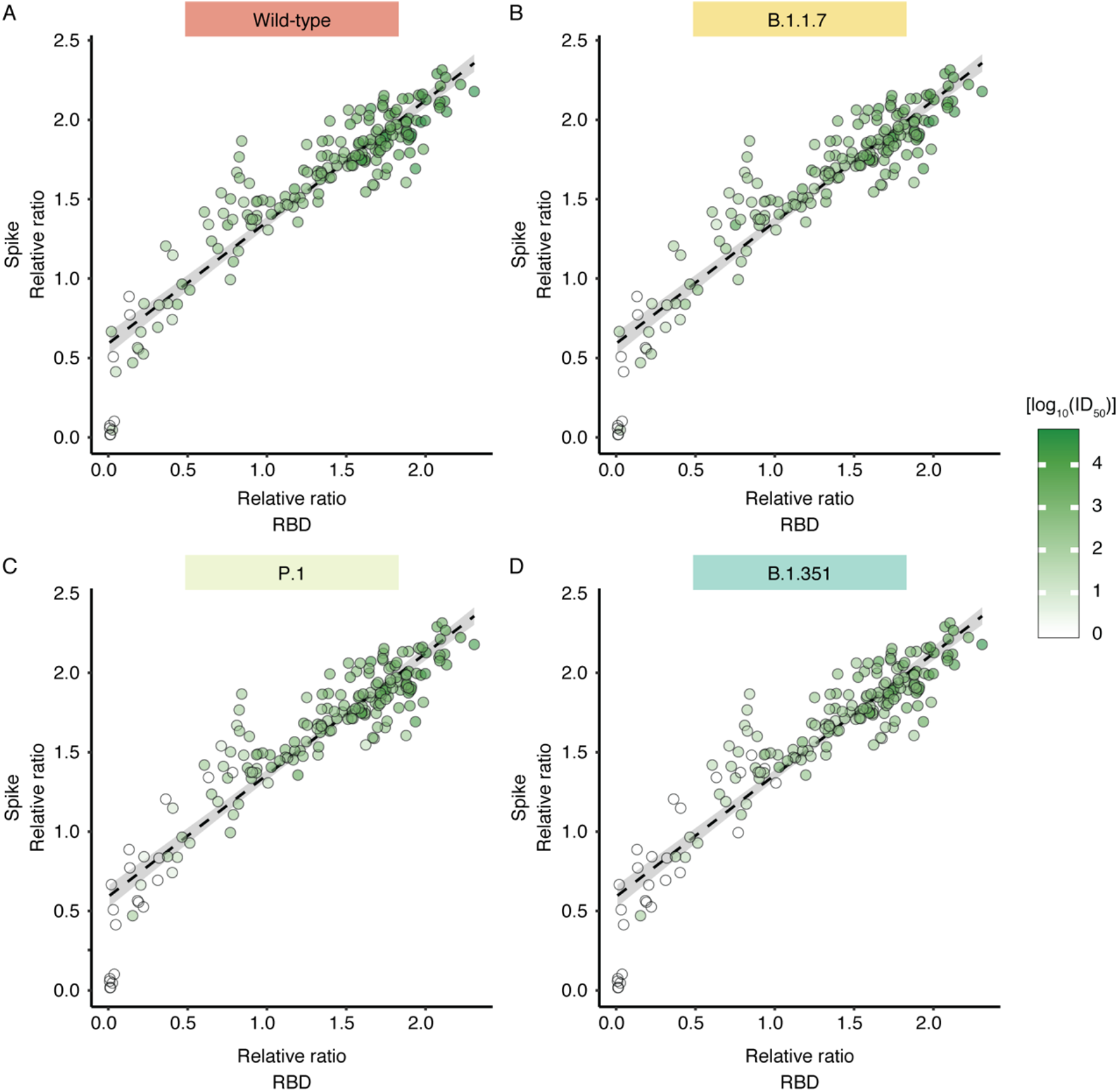
Correlation of neutralization of wild-type and VOCs based on spike and RBD IgG levels. Shades of green depict the degree of neutralization capacity (log_10_ID_50_). The dashed line indicates the regression line, and grey shading indicates the 95% confidence interval. For all plots, Spearman’s rho = 0.89, *P*-value < 0.001.

**Supplemental Figure 6.**
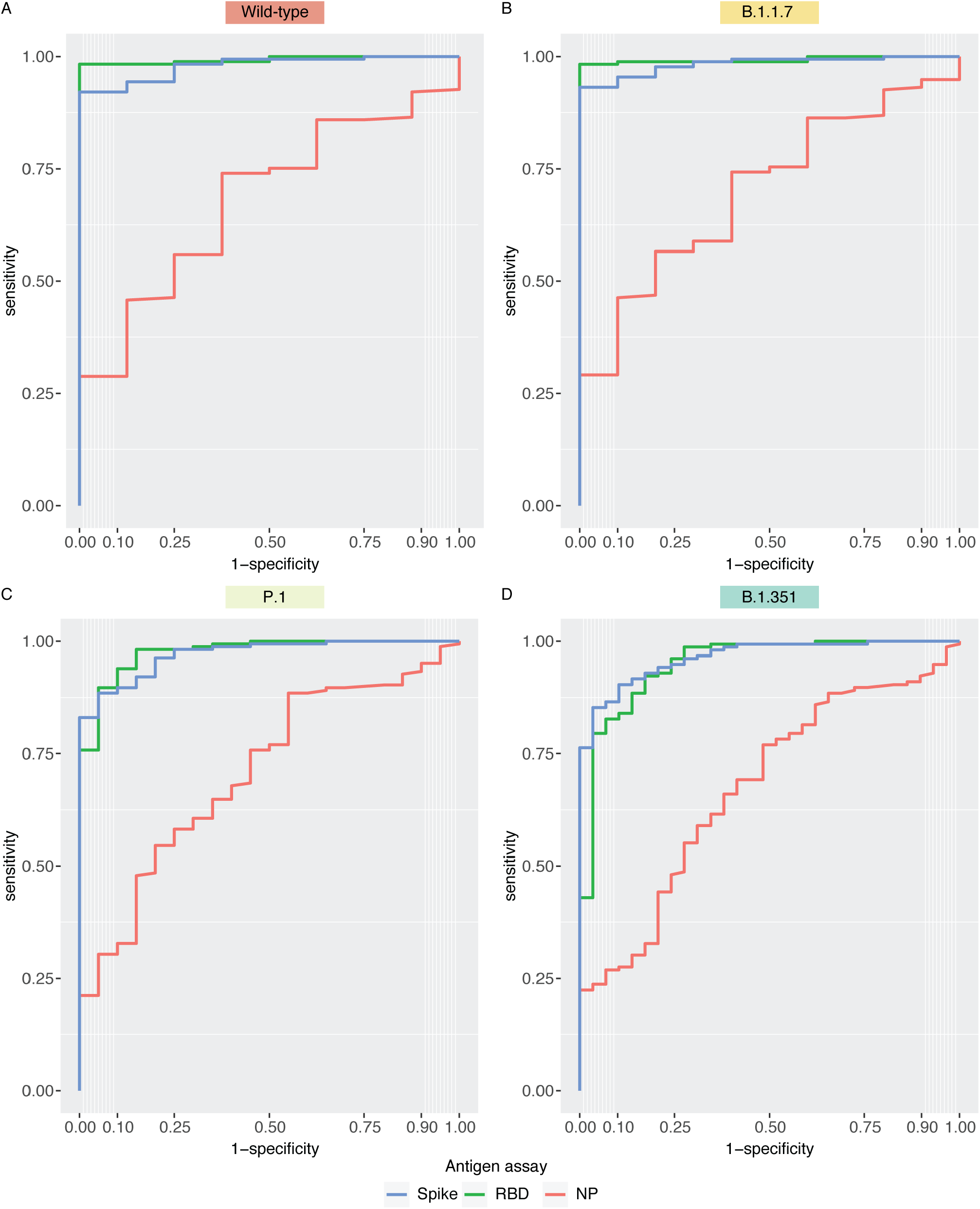
Receiver operating characteristic (ROC) analysis of ELISA-based assays across the VOCs tested with the VLP assay. Within each of the VOCs tested, an ID50 = 0 was classified as a negative response, and an ID50 > 0 was classified as a positive response.

## References

1. Hsu, A., et al. Report: Understanding the impact of COVID-19 on residents of Canada’s long-term care homes — ongoing challenges and policy responses. LTCcovid.org, International Long-Term Care Policy Network, CPEC-LSE, 2021.

2. Brown, K., et al. Early impact of Ontario’s COVID-19 vaccine rollout on long-term care home residents and health care workers. Science Briefs of the Ontario COVID-19 Science Advisory Table. 2021. 2.

3. Ontario, G.o., Managing long-term care home (LTCH) COVID-19 VOC outbreaks in the post-vaccine era, P.H. Ontario, Editor. 2021.

4. Williams, C., et al., COVID-19 Outbreak Associated with a SARS-CoV-2 P.1 Lineage in a Long-Term Care Home after Implementation of a Vaccination Program - Ontario, April-May 2021. Clin Infect Dis, 2021.

5. Vanker, A., et al., Adverse Outcomes Associated with SARS-CoV-2 variant B.1.351 Infection in Vaccinated Residents of a Long Term Care Home, Ontario, Canada. Clin Infect Dis, 2021.

6. Bailly, B., et al., BNT162b2 mRNA vaccination did not prevent an outbreak of SARS COV-2 variant 501Y.V2 in an elderly nursing home but reduced transmission and disease severity. Clin Infect Dis, 2021.

7. Orsi, A., et al., Outbreak of SARS-CoV-2 Lineage 20I/501Y.V1 in a Nursing Home Underlines the Crucial Role of Vaccination in Both Residents and Staff. Vaccines (Basel), 2021. 9(6).

8. 1. Canaday, D.H., et al., Reduced BNT162b2 mRNA vaccine response in SARS-CoV-2-naive nursing home residents. Clin Infect Dis, 2021.

9. Brockman, M.A., et al., Weak humoral immune reactivity among residents of long-term care facilities following one dose of the BNT162b2 mRNA COVID-19 vaccine. medRxiv, 2021.

10. Prendecki, M., et al., Effect of previous SARS-CoV-2 infection on humoral and T-cell responses to single-dose BNT162b2 vaccine. Lancet, 2021. 397(10280): p. 1178–1181.

11. Collier, D.A., et al., Age-related immune response heterogeneity to SARS-CoV-2 vaccine BNT162b2. Nature, 2021.

12. Moustsen-Helms, I.R., et al., Vaccine effectiveness after 1st and 2nd dose of the BNT162b2 mRNA Covid-19 Vaccine in long-term care facility residents and healthcare workers – a Danish cohort study. 2021, medRxiv.

13. Boyarsky, B.J., et al., Immunogenicity of a Single Dose of SARS-CoV-2 Messenger RNA Vaccine in Solid Organ Transplant Recipients. JAMA, 2021. 325(17): p. 1784–1786.

14. Boyarsky, B.J., et al., Antibody Response to 2-Dose SARS-CoV-2 mRNA Vaccine Series in Solid Organ Transplant Recipients. JAMA, 2021. 325(21): p. 2204–2206.

15. Montoya, J.G., et al., Differences in IgG antibody responses following BNT162b2 and mRNA-1273 Vaccines. bioRxiv, 2021: p. 2021.06.18.449086.

16. Khoury, D.S., et al., Neutralizing antibody levels are highly predictive of immune protection from symptomatic SARS-CoV-2 infection. Nat Med, 2021. 27(7): p. 1205–1211.

17. de Jong, J.C., et al., Haemagglutination-inhibiting antibody to influenza virus. Dev Biol (Basel), 2003. 115: p. 63–73.

18. Planas, D., et al., Sensitivity of infectious SARS-CoV-2 B.1.1.7 and B.1.351 variants to neutralizing antibodies. Nat Med, 2021. 27(5): p. 917–924.

19. Liu, C., et al., Reduced neutralization of SARS-CoV-2 B.1.617 by vaccine and convalescent serum. Cell, 2021.

20. Dejnirattisai, W., et al., Antibody evasion by the P.1 strain of SARS-CoV-2. Cell, 2021. 184(11): p. 2939–2954 e9.

21. Kustin, T., et al., Evidence for increased breakthrough rates of SARS-CoV-2 variants of concern in BNT162b2-mRNA-vaccinated individuals. Nat Med, 2021.

22. Wall, E.C., et al., Neutralising antibody activity against SARS-CoV-2 VOCs B.1.617.2 and B.1.351 by BNT162b2 vaccination. Lancet, 2021. 397(10292): p. 2331–2333.

23. Isho, B., et al., Persistence of serum and saliva antibody responses to SARS-CoV-2 spike antigens in COVID-19 patients. Sci Immunol, 2020. 5(52).

24. Crawford, K.H.D., et al., Protocol and Reagents for Pseudotyping Lentiviral Particles with SARS-CoV-2 Spike Protein for Neutralization Assays. Viruses, 2020. 12(5).

25. Abe, K.T., et al., A simple protein-based surrogate neutralization assay for SARS-CoV-2. JCI Insight, 2020. 5(19).

26. Liu, J., et al., Preclinical evaluation of a SARS-CoV-2 mRNA vaccine PTX-COVID19-B. bioRxiv, 2021: p. 2021.05.11.443286.

27. Harvey, W.T., et al., SARS-CoV-2 variants, spike mutations and immune escape. Nat Rev Microbiol, 2021. 19(7): p. 409–424.

28. Gupta, R.K., Will SARS-CoV-2 variants of concern affect the promise of vaccines? Nat Rev Immunol, 2021. 21(6): p. 340–341.

29. Breznik, J.A., et al., Antibody Responses 3-5 Months Post-Vaccination with mRNA-1273 or BNT163b2 in Nursing Home Residents. medRxiv, 2021: p. 2021.08.17.21262152.

30. Sette, A. and S. Crotty, Adaptive immunity to SARS-CoV-2 and COVID-19. Cell, 2021. 184(4): p. 861–880.

31. Polack, F.P., et al., Safety and Efficacy of the BNT162b2 mRNA Covid-19 Vaccine. N Engl J Med, 2020. 383(27): p. 2603–2615.

32. Baden, L.R., et al., Efficacy and Safety of the mRNA-1273 SARS-CoV-2 Vaccine. N Engl J Med, 2021. 384(5): p. 403–416.

33. Yau, K., et al., The Humoral Response to the BNT162b2 Vaccine in Hemodialysis Patients. medRxiv, 2021: p. 2021.05.24.21257425.

34. Sheikh-Mohamed, S., et al., A mucosal antibody response is induced by intra-muscular SARS-CoV-2 mRNA vaccination. medRxiv, 2021: p. 2021.08.01.21261297.

35. Mendoza, E.J., et al., Two Detailed Plaque Assay Protocols for the Quantification of Infectious SARS-CoV-2. Curr Protoc Microbiol, 2020. 57(1): p. ecpmc105.

36. Wang, S., et al., Assays for the assessment of neutralizing antibody activities against Severe Acute Respiratory Syndrome (SARS) associated coronavirus (SCV). J Immunol Methods, 2005. 301(1-2): p. 21–30.

37. Shigayeva, A., et al., Invasive Pneumococcal Disease Among Immunocompromised Persons: Implications for Vaccination Programs. Clin Infect Dis, 2016. 62(2): p. 139–47.

